# Longitudinal Relationships Between Cannabis and Tobacco Use and Symptom Severity in Individuals at Clinical High Risk for Psychosis

**DOI:** 10.64898/2026.03.16.26347411

**Authors:** Yunong Bai, Simon Vandekar, Brandee Feola, Jean Addington, Carrie E. Bearden, Kristin Cadenhead, Tyrone D. Cannon, Barbara Cornblatt, Matcheri Keshavan, Daniel H. Mathalon, Diana O. Perkins, Larry Seidman, William S. Stone, Ming T. Tsuang, Elaine F. Walker, Scott Woods, Ricardo E Carrión, Heather Burrell Ward

**Affiliations:** Department of Psychiatry and Behavioral Sciences, Vanderbilt University Medical Center, Nashville, TN, USA; Department of Biostatistics, Vanderbilt University Medical Center, Nashville, TN, USA; Department of Psychiatry, Hotchkiss Brain Institute, University of Calgary, Alberta, Canada; Semel Institute for Neuroscience and Human Behavior, Departments of Psychiatry and Behavioral Sciences and Psychology, University of California, Los Angeles, Los Angeles, CA, USA; Department of Psychiatry, University of California, San Diego, La Jolla, CA, USA; Department of Psychology and Psychiatry, Yale University, New Haven, CT, USA; Institute for Behavioral Science, Feinstein Institutes for Medical Research, Northwell Health System, Manhasset, NY, USA; Department of Psychiatry, Donald and Barbara Zucker School of Medicine at Hofstra/Northwell, Hempstead, New York, NY, USA; Department of Psychiatry, Beth Israel Deaconess Medical Center and Harvard Medical School, Boston, MA, USA; Department of Psychiatry and Behavioral Sciences, University of California, San Francisco, San Francisco, CA, USA; Veterans Affairs San Francisco Health Care System, San Francisco, CA, USA; Department of Psychiatry, University of North Carolina at Chapel Hill, Chapel Hill, NC, USA; Department of Psychology, Emory University, Atlanta, GA, USA; Department of Psychiatry, Yale University, New Haven, CT, USA

**Keywords:** clinical high risk for psychosis, psychosis, cannabis, tobacco, longitudinal analysis

## Abstract

**Objective:** Tobacco and cannabis are the most used substances among individuals at clinical high risk for psychosis (CHR-P), but it remains controversial whether substance use drives symptom exacerbation and psychosis transition, or vice versa. We investigated longitudinal dose-response relationships of tobacco and cannabis use with clinical presentation in a CHR-P population.

**Methods:** Data was obtained from the North American Prodrome Longitudinal Study (NAPLS2) CHR-P cohort (n=764). Participants were assessed every 6 months over two years. Substance use frequency, psychiatric symptoms (psychosis, depression, anxiety, and social anxiety), global social and role functioning, and neurocognitive performance were measured. Linear mixed effect models were used to model the relationship between substance use and clinical measurements across visits, and that between baseline use and trajectory of symptoms, functioning, and cognition.

**Results:** Psychiatric symptoms, functioning, and cognitive performance improved, while tobacco and cannabis use frequency did not change over two years for CHR-P individuals in NAPLS2. Heavier tobacco and cannabis use at current visit predicted worse anxiety at next visit (tobacco: β=0.178, p=0.033; cannabis: β=0.162, p=0.018). Better social functioning predicted heavier tobacco (β=0.178, p<0.001) and cannabis: (β=0.162, p<0.001) use at next visit. We observed a significant baseline cannabis-by-time interaction, where heavier baseline cannabis use predicted slower improvement of negative symptoms (β=0.159, p=0.0017, FDRp=0.0067) and deterioration of role function (β=-0.046, p=0.018).

**Conclusions:** In CHR-R, current tobacco and cannabis use predicted worse anxiety at future visits. Baseline cannabis use frequency predicts worse clinical trajectory, especially for negative symptoms.

## Introduction

Substance use is highly prevalent among people at clinical high risk for psychosis (CHR-P). Tobacco and cannabis use are most common, with an estimated prevalence of up to 64% of CHR-P using tobacco (1) and 48.7% using cannabis (2). An important, but unresolved question is whether substance use worsens symptoms and drives the transition to psychosis, or if it represents a form of self-medication.

In the general population, both tobacco and cannabis use have been associated with psychosis. Prospective population-based cohort studies found that tobacco smoking was associated with a higher risk of subsequent schizophrenia (SZ) (3) with a dose-response relationship (4). Cannabis use may lead to transient psychotic symptoms in the general population (5) and negatively impacts cognitive function, especially if used starts in adolescence (6).

Similar trends between tobacco/cannabis use and clinical presentation extend to patients with SZ. Cannabis use has been consistently shown to predict worse clinical outcomes in people with schizophrenia spectrum disorders (7). Among patients with first-episode psychosis, a retrospective study found a higher risk of developing psychosis in those with a recent increase in tobacco and cannabis use (8). One longitudinal study by Penzel et al. demonstrated that among first-episode psychosis patients who use cannabis, continued cannabis use was related to slower recovery of psychotic symptoms and functioning over 18 months compared to those with complete abstinence (9).

However, in the CHR-P population, evidence has not converged regarding the relationships between tobacco/cannabis use and psychiatric presentation. Regarding transition to psychosis, several cross-sectional studies on CHR-P individuals reported no association between baseline tobacco or cannabis use and transition (10,11), and a meta-analysis done by Farris et al (2) found no link between cannabis use and higher risk of transition in CHR-P. Regarding psychiatric symptoms irrespective of transition, cross-sectional studies report mixed findings regarding associations between cannabis use and psychiatric symptoms (12–16). In the same longitudinal study by Penzel et al., CHR-P with continued cannabis use did not differ in clinical trajectory compared to those with complete abstinence (9).

Beyond psychiatric symptoms, previous literature also showed conflicting findings regarding tobacco/cannabis use and cognitive performance. Cognitive deficits precede the onset of psychotic symptoms in CHR-P (17), and CHR-P individuals often exhibit significant functional impairment (18). However, paradoxical cognitive impacts have been reported for both tobacco and cannabis use in CHR-P. Even though cognitive performance was worse in CHR-P compared to demographically-matched controls (19), within the CHR-P group, tobacco use was associated with better cognition (20), and occasional cannabis use was associated with higher premorbid functioning and intelligence (21). Yet despite these cross-sectional findings, acute administration of tetrahydrocannabinol (THC) worsened cognitive performance in CHR-P (22).

In short, the field has not converged on this debate: in CHR-P individuals, does tobacco or cannabis use worsen overall clinical presentation, or is tobacco or cannabis use an attempt to self-medicate their unmet psychiatric, cognitive, and functional needs? Answering these questions can guide early intervention. However, selecting an appropriate study design for these questions is challenging: whereas cross-sectional studies cannot elucidate causal relationships between substance use and symptoms, interventional studies in this population (i.e., administering cannabis to CHR-P adolescents) pose an ethical dilemma. Therefore, analysis of longitudinal data offers an ideal method to understand directionality of substance use and symptom relationships.

We therefore analyzed data from the North American Prodrome Longitudinal Study (NAPLS2), a longitudinal dataset following a cohort of CHR-P individuals over two years with repeated assessments of substance use, psychiatric symptoms, cognitive performance, and functioning. While many previous studies have treated substance use as a binary variable (use vs. no use, (9)), NAPLS2 assessed frequency of substance use over the past month, allowing assessment of dose-response relationships between substance use and symptom severity.

To disentangle temporal associations between substance use and clinical presentation of psychiatric symptoms, functioning, and cognitive performance, we asked 3 key questions: 1) Does current substance use predict clinical presentation at the next visit? 2) Does current clinical presentation predict substance use at the next visit? 3) Does baseline substance use predict the trajectory of symptoms, functioning, and cognition? (Figure 1) Based on the competing causal and self-medication models, we proposed three sets of hypotheses: 1) Current cannabis use would predict increased positive symptoms at the next visit; 2) current psychotic symptoms would predict increased tobacco use at the next visit; and 3) more frequent tobacco and cannabis use at baseline would be associated with a worse trajectory of psychiatric symptoms, functioning, and cognitive performance over the study period.

**Figure 1.**
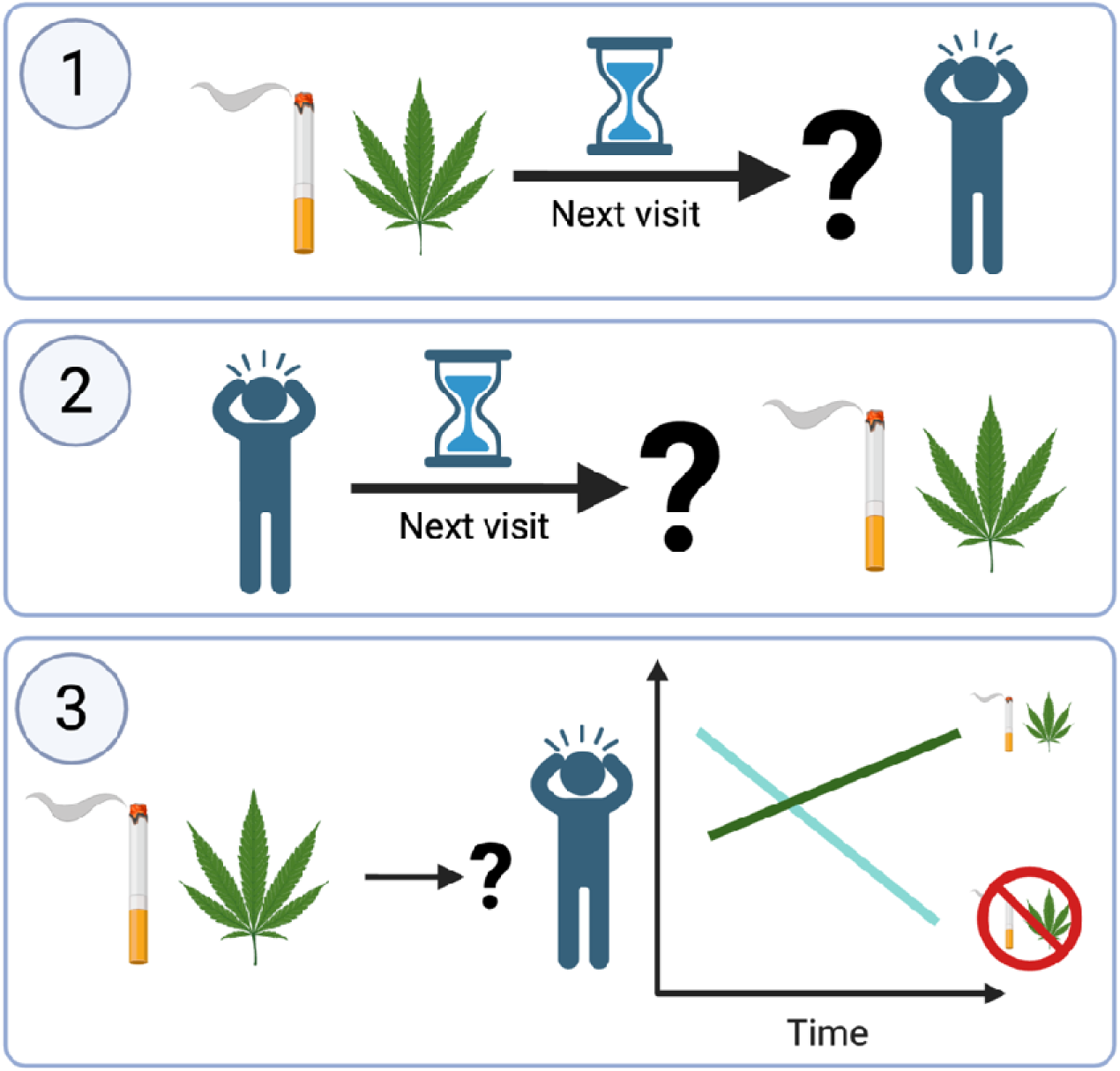
Graphical Abstract. We used data from the North American Prodrome Longitudinal Study 2 (NAPLS2) to understand longitudinal relationships between substance use and clinical presentation of psychiatric symptoms, functioning, and cognitive performance in individuals at clinical high risk for psychosis. To do this, we asked 3 key questions: 1) Does current substance use predict clinical presentation at the next visit? 2) Does current clinical presentation predict substance use at the next visit? and 3) Does baseline substance use predict the trajectory of symptoms, functioning, and cognition?

## Methods

### Participants

NAPLS2 is a longitudinal case-control study of individuals at CHR-P for psychosis across 8 sites in North America. Individuals meeting CHR-P criteria (N=764) and healthy controls (N=280) were enrolled. Only participants meeting CHR-P criteria were included in this analysis. Participants underwent assessment at baseline and every 6 months for two years and upon conversion to psychosis (if applicable) from January 2009 to April 2013. Data from conversion visits were excluded from this analysis for consistency in modeling time intervals in longitudinal analyses. See supplement for details.

### Measures

#### Clinical

The Structured Clinical Interview for DSM was used to exclude psychosis and to identify DSM-IV Axis I or cluster A personality disorders. We categorized DSM-IV disorders into any affective disorders or anxiety disorders. Conversion to psychosis was based on the Structured Interview for Psychosis-Risk Syndromes (23), defined as meeting the Presence of Psychosis Syndrome criteria. See Supplement for details.

#### Substance use

Current substance use at baseline was measured using the Alcohol Use Scale/Drug Use Scale (AUS/DUS) (24), which assesses substance use frequency over the past 30 days on an ordinal scale. Tobacco use was rated in cigarettes per day (0 = no use, 1 = occasionally, 2 = <10 per day, 3 = 11-25 per day, 4 = >25 per day). Cannabis frequency was reported as (0 = no use, 1 = once or twice per month, 2 = 3-4 times per month, 3 = 1-2 times per week, 4 = 3-4 times per week, 5 = almost daily). See Supplement for details.

#### Psychiatric Symptoms

Severity of psychosis-risk symptoms at baseline was rated using the Scale of Psychosis-Risk Symptoms (SOPS) (23); scoring included SOPS total score (“total psychosis-risk symptoms”), positive symptom subscore (“positive symptoms”), negative symptom subscore (“negative symptoms”), disorganization symptom subscore (“disorganization symptoms”), and general symptom subscore (“general symptoms”). Anxiety was measured with the Self-Rating Anxiety Scale (SAS, “anxiety”) (25). Social anxiety was measured with Social Interaction Anxiety Scale (SIAS, “social anxiety”) (26). Depressive symptoms were assessed using the Calgary Depression Scale for Schizophrenia (CDSS, “depression”) (27). See Supplement for details.

#### Global Functioning

Functioning was measured using the clinician-rated Global Functioning: Social and Role Scales (GFS and GFR), which assess intimate relationships, family involvement, peer relationships, and peer conflict (Social) and performance in specific roles like school or work (Role).

#### Cognitive Performance

Neurocognition was assessed with six domains from the MATRICS Consensus Cognitive Battery (MCCB) (28–30), including Processing Speed, Attention/Vigilance, Working Memory, Verbal Learning, Visual Learning, and Reasoning and Problem Solving. See Supplement for details.

### Statistical Approach

#### Within-Visit Associations

We used linear mixed effect (LME) models with subject-level random intercepts to assess the association between tobacco/cannabis use frequency and psychiatric symptoms, functioning, and cognition at each visit. See Supplement for details.

#### Symptoms and Substance Use Trajectory

Random intercept LME models were used to model the longitudinal trajectory of total psychosis-risk symptoms, social functioning, role functioning, and cognition over the study period. Ordinal regression models (ORMs) were used to model the longitudinal trajectory of tobacco and cannabis use frequency. Visit number, modeled as a continuous variable, was used as the explanatory variable.

#### Predict Future Symptom Severity Using Current Substance Use

Random intercept LME models were used to predict symptoms at the next visit using current tobacco or cannabis use frequency as the explanatory variable, and current symptoms, age at visit, sex, and antipsychotics medication dosage (chlorpromazine equivalent) as covariates. Substance use frequency was modeled as a continuous variable. Symptoms included total psychosis-risk and subscores (positive, negative, disorganization, general), anxiety, social anxiety, depression, social functioning, role functioning, global cognition and cognitive domain subscores. False discovery rate (FDR) adjusted p-values were computed separately across psychosis symptom subscores and across cognitive subdomain scores.

#### Predict Future Substance Use Using Current Symptom Severity

Ordinal regression models (ORMs) were used to predict tobacco or cannabis use frequency at the next visit using individual symptom and tobacco or cannabis use at current visit. Individual symptoms as listed above were used as the explanatory variable. Current substance use, age at visit, sex, and antipsychotics dosage were covariates. FDR adjusted p-values were computed separately across psychosis symptom subscores and across cognitive subdomain scores.

#### Impact of Baseline Substance Use on Symptom Trajectory

Random intercept LME models were used to model symptoms including baseline substance use frequency, visit number, and their interaction as explanatory variables. Sex and antipsychotic dosage were covariates. FDR adjusted p-values were computed separately across psychosis symptom subscores and across cognitive subdomain scores. Since our finding on baseline cannabis use and positive symptom trajectory contradicted our hypothesis, we conducted two post-hoc analyses to investigate the relationship between baseline cannabis use and positive symptom severity both at baseline and throughout the study period: 1) using a generalized linear model, we modeled baseline positive symptom severity using baseline cannabis use frequency, controlling for age at visit, sex, and antipsychotics dosage; 2) using a random intercept LME model, we modeled positive symptom severity at each visit using baseline cannabis use, controlling for the same covariates.

Predictor significance in all LME models was assessed using type II analysis of variance (ANOVA). Cluster robust covariance matrix estimation was used with ORM to account for repeated measurements within each subject. Separate models were built for each outcome variable of interest in the trajectory analysis, and for each pair of substance and symptom in other analyses.

## Results

A total of 764 CHR-P individuals were enrolled in NAPLS2. Average age at baseline was 18.50 (SD=4.23). Only 7.46% of CHR-P individuals were taking antipsychotics at baseline. Approximately one-third of participants had a baseline diagnosis of an affective disorder (38.48%) or anxiety disorder (39.79%). Data completion rate for substance use and clinical measurements declined over the study period, ranging from between 89.92% and 98.56% for the baseline visit, to between 30.72% and 36.51% for the 24-month follow-up visit. Most CHR-P participants reported no use of tobacco or cannabis across all visits (Figure S1). See Table S1 for details regarding data completion, tobacco and cannabis use frequency, and average severity for all symptoms, functioning, and cognition of interest. Within each visit, heavier tobacco and cannabis use was significantly associated with more severe psychiatric symptoms; See Supplement for details.

### Symptoms and Substance Use Trajectory: Symptoms improved while substance use did not change for CHR-P individuals over two years

We examined whether symptoms (psychosis-risk, anxiety, depression), functioning, cognition, and substance use changed over two years. Visit number significantly predicted total psychosis-risk symptoms (β=-3.804, p<0.001), anxiety (β=-2.204, p<0.001), depression (β=-0.750, p<0.001), role functioning (β=0.082, p=0.002), social functioning (β=0.173, p<0.001), and global cognition (β=0.058, p<0.001), indicating improvement in all four measurements over two years (Figure S2, Table S2). Tobacco and cannabis use frequency did not change over two years (Figure S2, Table S2).

### Current Substance Use Predicts Symptom Severity at Next Visit: Heavier tobacco and cannabis use predict higher anxiety at next visit

We assessed whether substance use at the current visit predicted symptoms, functioning, and cognitive performance at the next visit when controlling for current clinical presentation. More frequent current tobacco use predicted worse anxiety at next visit (β=0.712, p=0.033; Figure 2A, Table S4). More frequent current cannabis use predicted worse anxiety (β=0.511, p=0.018; Figure 2B, Table S4) and better social functioning (β=0.060, p=0.009; Figure 2C, Table S4) at next visit. Current tobacco or cannabis use frequency did not predict psychosis symptoms (total and domain subscores) or depression symptoms, role functioning, or cognition (Table S4), suggesting that in the short-term, the primary impact of substanc use is on anxiety rather than on positive or negative symptoms.

**Figure 2.**
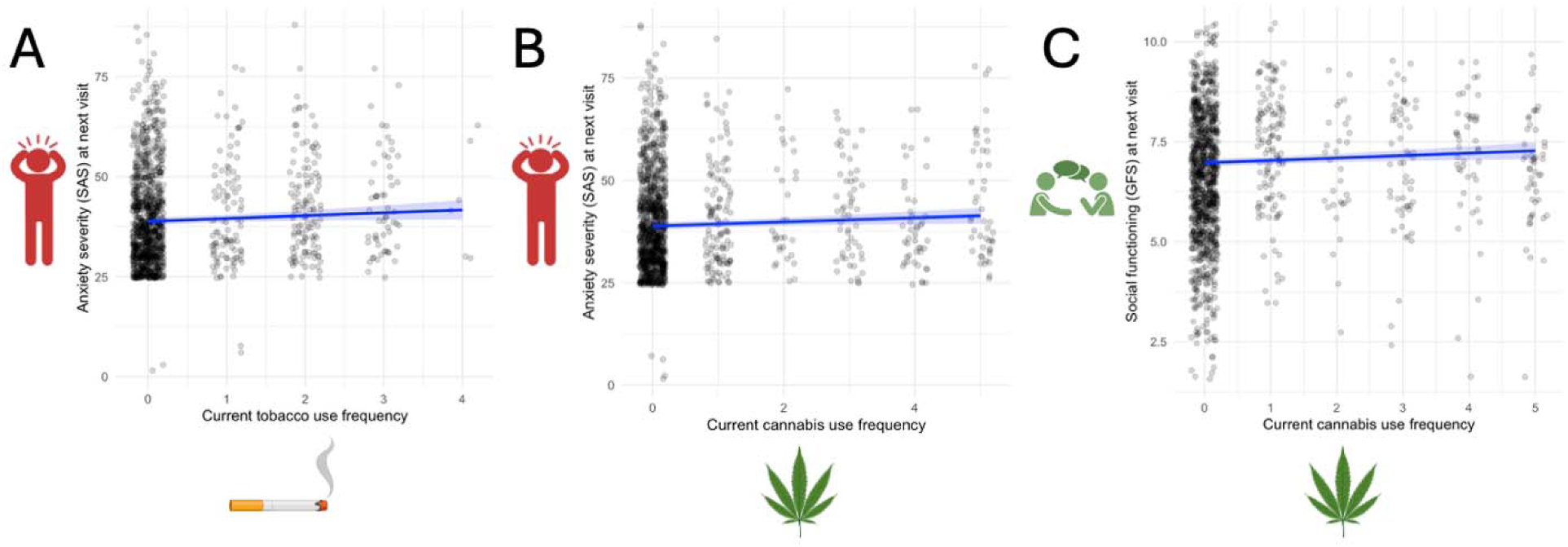
Current Substance Use Predicts Clinical Presentation at Next Visit. Heavier tobacco use predicts more severe anxiety symptoms (A) at next visit. Heavier cannabis use predicts more severe anxiety symptoms (B) and better social functioning (C) at next visit. GFS, Global Functioning: Social Scale; SAS, Self-Rating Anxiety Scale.

### Current Symptoms Predict Substance Use at Next Visit: Better social functioning predicts heavier tobacco and cannabis use at next visit

We then asked whether current symptoms, functioning, and cognitive performance predicted substance use at the next visit, controlling for current substance use. Better social functioning predicted both heavier tobacco use (β=0.178, p<0.001; Figure 3A, Table S5) and heavier cannabis use (β=0.162, p<0.001; Figure 3B, Table S5) at next visit. Current symptoms, role functioning, and cognitive measurement did not predict tobacco or cannabis use at next visit (Table S5).

**Figure 3.**
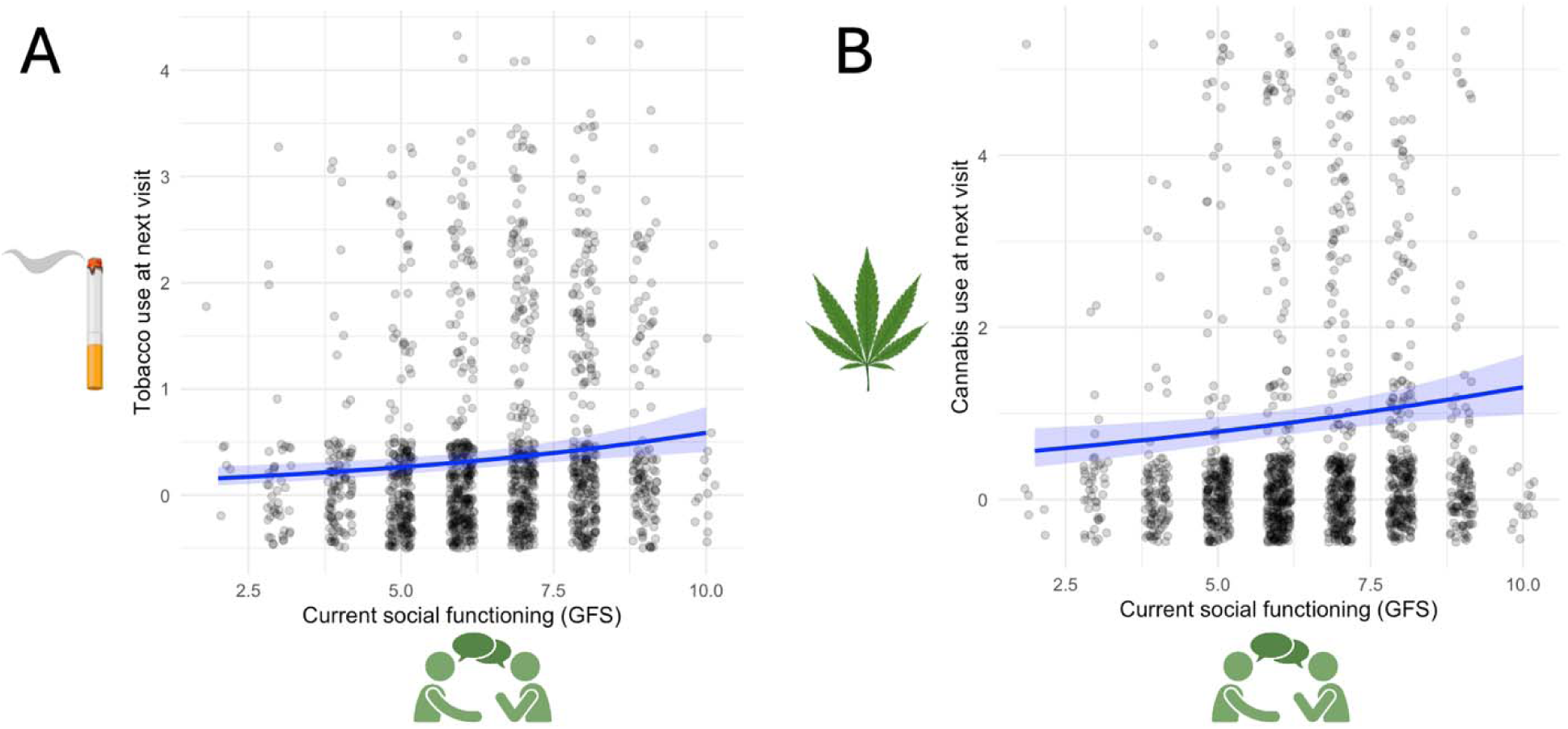
Current Social Functioning Predicts Substance Use at Next Visit. Better social functioning predicts heavier tobacco use (A) and heavier cannabis use (B) at next visit, when controlling for current substance use. GFS, Global Functioning: Social Scale.

### Impact of Baseline Substance Use on Clinical Trajectory: Heavier baseline cannabis use predicted slower recovery of negative symptoms and deterioration of role functioning in CHR-P individuals

We investigated if baseline substance use impacted trajectory of symptoms, functioning, and cognition. We observed significant baseline cannabis-by-time interaction, where heavier baseline cannabis us predicted slower recovery of negative symptoms (β=0.159, p=0.0017, FDRp=0.0067; Figure 4A), deterioration of role functioning (β=-0.0046, p=0.018; Figure 4B), and deterioration of visual learning (β=-0.031, p=0.00657, FDRp=0.046; Figure 4C, Table S6) but did not significantly modify trajectory of other symptoms, functioning, or cognition (Table S6). Baseline tobacco-by-time interaction was observed for social functioning, where heavier baseline tobacco use predicted slower improvement of social functioning (β=-0.033, p=0.049; Figure S3, Table S6). Baseline tobacco use frequency did not modify other clinical trajectories (Table S6).

**Figure 4.**
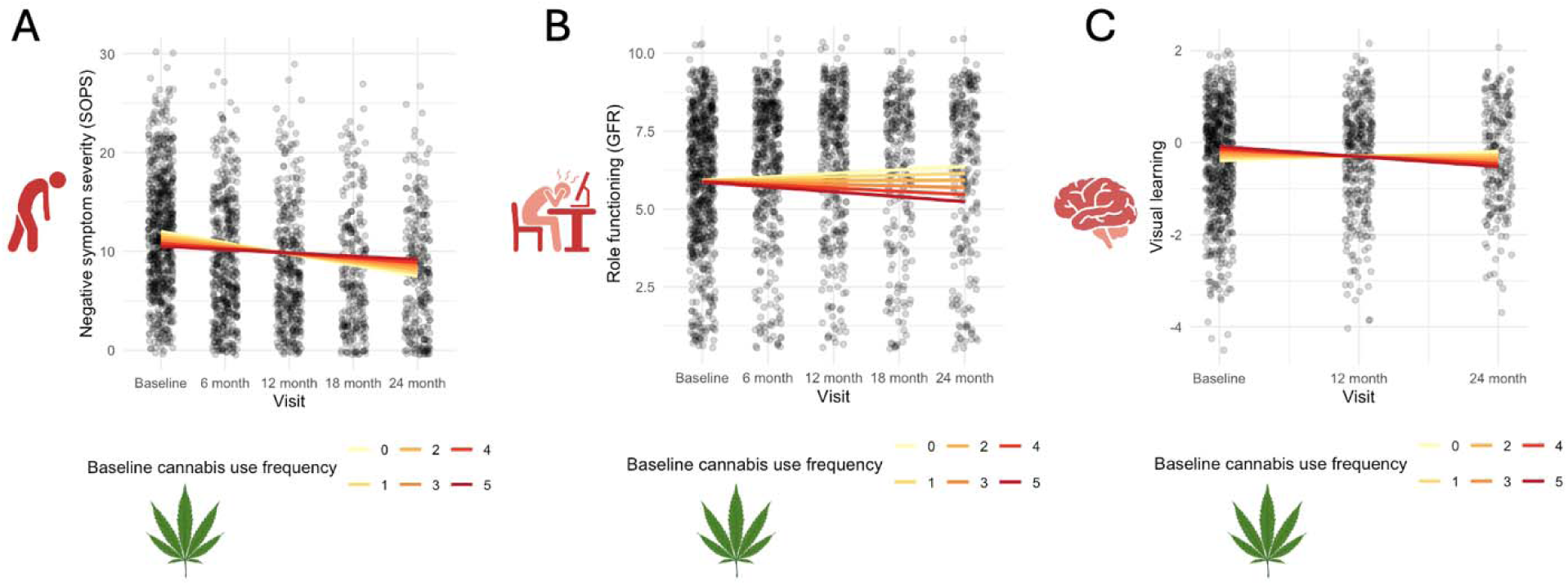
Impact of Baseline Substance Use on Symptom Trajectory. Heavier baseline cannabis use predicts (A) slower recovery of general symptoms, (B) deterioration of role functioning, and (C) deterioration of visual learning. GFR, Global Functioning: Role Scale; SOPS, Scale of Psychosis-Risk Symptoms.

Given baseline cannabis use did not impact positive symptom trajectory, contradicting our hypotheses, we performed post-hoc analyses. We found that heavier baseline cannabis use was associated with wors positive symptoms both at baseline (β=0.263, p=0.020; Figure S4) and at each visit (β=0.359, p=0.003; Figure S4), though the overall trajectory did not change.

## Discussion

In this large, longitudinal study of CHR-P individuals, we examined the complex temporal relationships between substance use and clinical outcomes. Applying lead-lag analyses to disentangle the temporal precedence of risk factors and outcomes, we addressed whether substance use drives clinical deterioration or if emerging symptoms lead to substance use (i.e., self-medication). Our results reveal a complex picture with three key findings. First, heavier tobacco and cannabis use both forecasted anxiety exacerbation. Second, we found a bidirectional relationship between social functioning and cannabis use. Better social functioning predicted heavier future substance use, while cannabis use also predicted better future social functioning. Third, heavier baseline cannabis use was a prognostic marker for poorer long-term trajectories for negative symptoms, role functioning, and visual learning (Figure 5).

**Figure 5.**
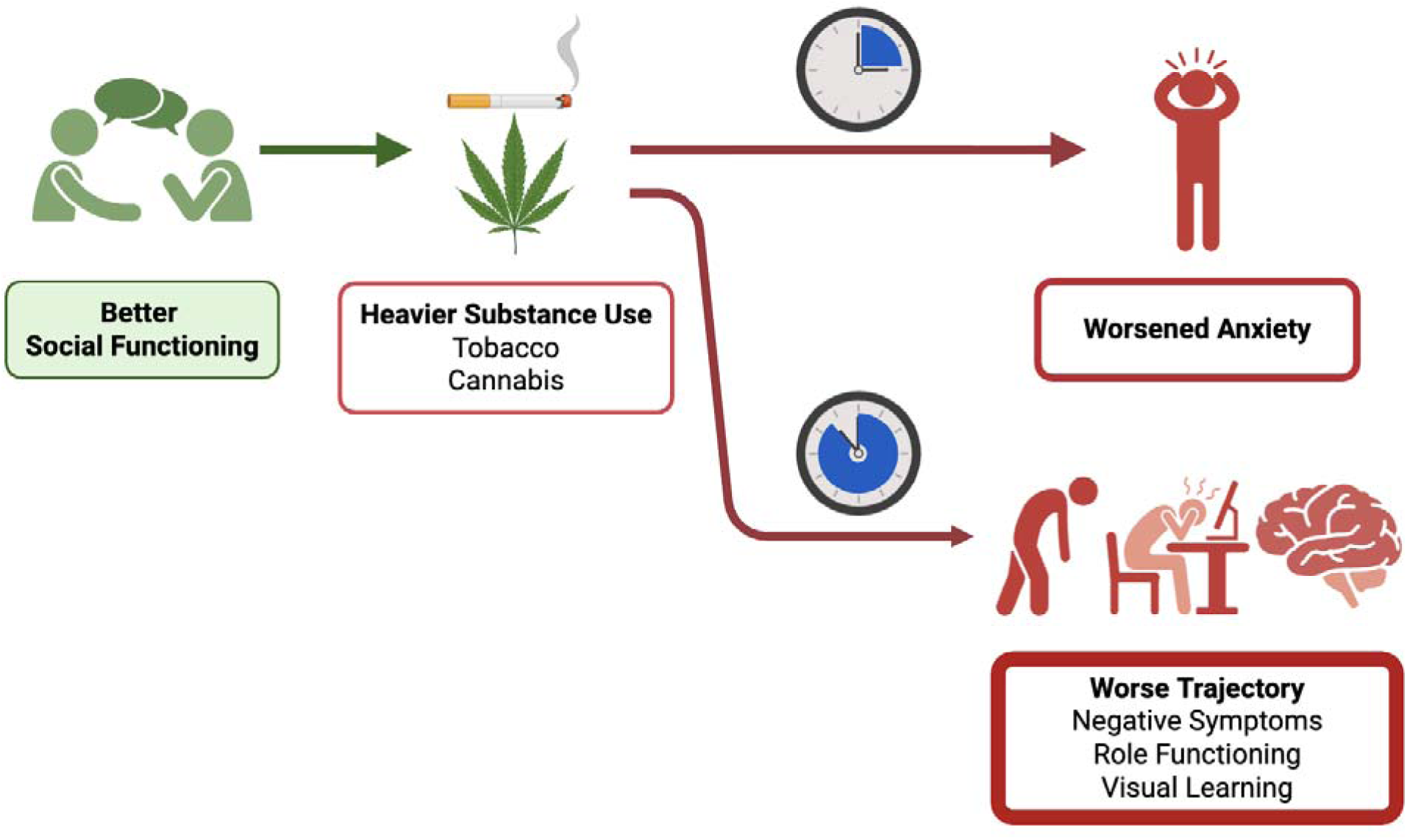
Proposed Model Relationships Between Substance Use and Psychiatric Symptoms in CHR-P Individuals. The results of our longitudinal analyses suggest that social functioning facilitate tobacco and cannabis use, while substance use worsens anxiety symptoms. Heavier baseline cannabis use prognosticates worse recovery trajectory of negative symptoms, role functioning, and cognition.

Our longitudinal findings revealed a critical temporal relationship between tobacco/cannabis use and anxiety in CHR-P individuals. Although anxiety is not a diagnostic factor for CHR-P, anxiety symptoms and disorders are commonly experienced across the stages of psychosis development (31). Critically, anxiety symptoms are associated with worse clinical outcomes and functioning in CHR-P (32). Anxiety and substance use disorders are also highly comorbid (33). Therefore, anxiety in CHR-P is a key factor to consider when examining substance use in this population. Our findings demonstrated both short- and long-term effects, where tobacco and cannabis use in CHR-P youths increase their anxiety symptoms in the short-term, and exacerbate the long-term trajectory of other symptoms. Early interventions should break this cycle by simultaneously targeting both anxiety and substance use.

Our results did not support self-medication hypothesis, as we found that higher levels of anxiety, depression, or psychosis-risk symptoms did not predict an escalation in substance use at the next visit. However, our analysis revealed a complex, bidirectional relationship between social functioning and substance use. In the short-term, more frequent cannabis use predicted better social functioning at the next visit. Conversely, better social functioning at a given visit predicted heavier future use of both cannabis and tobacco. This latter finding is consistent with and extends prior longitudinal work showing that CHR-P individuals who use cannabis tend to have higher social functioning than non-users (34). The lead-lag analysis in the present study builds upon this by establishing a specific temporal sequence, strengthening the hypothesis that greater social functioning facilitates future substance use. This presents a clinical paradox: while strengthening social connections is a typical therapeutic goal for CHR youths, this intervention may inadvertently create opportunities for substance use. Psychosocial interventions should promote social connection while preparing CHR-P individuals with skills and strategies for navigating social interactions that are pro-substance use, and it is crucial to also monitor for changes in substance use.

The complexity of this social dynamic is further underscored by the lack of a predictive relationship with the feeling of social anxiety itself. Social anxiety is the most common anxiety disorder in people at CHR-P, with a prevalence of approximately 50% (36). Social anxiety is associated with worse positive symptoms and functional outcomes in this population (37). One potential explanation for our findings is that the measure of anxiety (SAS) captures anxiety over the past few days, while the measure of social anxiety (SIAS) captures social anxiety in general and is not time-specific. Therefore, the social anxiety measure in the study may not be well suited to capture anxiety symptoms over time. Another possibility is that substance use may dampen perceived social anxiety, which is consistent with Gill et al’s interpretation on CHR-P reporting reward/recreational motivations for cannabis use (12). Our findings are also consistent with Reeves et al, who reported that trait anxiety, instead of social anxiety, mediated the relationship between cannabis use and positive symptoms in young adults in the general population (38). Therefore, it will be critical for future studies to investigate the associations between specific aspects of anxiety and types of substance use in CHR-P.

Furthermore, we found that heavier baseline cannabis use had deleterious effect on the recovery trajectory of negative symptoms, role functioning, and visual learning, but not that of positive symptoms. It is unsurprising that impaired recovery of negative symptoms tracked with further deterioration of role functioning, as the Global Functioning: Role scale (GFR) assesses performance in school or work, which is directly impacted by negative symptoms (39). Even though we did not observe a relationship between baseline cannabis use and trajectory of positive symptoms, our post-hoc analysis demonstrated that participants with heavier baseline cannabis use had persistently higher positive symptoms for the entire study duration. This implies that cannabis use is stably associated with positive symptoms in CHR-P, but baseline cannabis use failed to *alter* positive symptom trajectory, as positive symptoms were already substantially elevated at enrollment. In this light, heavier baseline cannabis use may be clinically applicable as a prognostic marker for worse recovery trajectory in CHR-P. Our findings of a longitudinal, dose-response relationship support a causal link between cannabis use and clinical deterioration in CHR-P, though it is also possible that other latent factors associated with baseline cannabis use are mechanically causing this deterioration, such as lower socioeconomic status or trauma exposure. Future studies should aim to disentangle the associations between cannabis use and long-term clinical outcomes.

Our study has several key strengths. First, this is the first longitudinal analysis of the relationships between tobacco and cannabis use and clinical presentation in CHR-P. Furthermore, our use of lead-lag longitudinal models allowed us to test for temporal precedence between substance use and clinical presentation, a critical advance over cross-sectional designs. Second, we identified dose-response relationships by measuring substance use frequency, as opposed to modeling substance use as a binary variable. Finally, we used a large sample of CHR-P individuals from multiple sites across North America who had adequate follow-up to support longitudinal statistical analysis.

There are several limitations to our analysis. Despite large sample size in the initial visits, we had significant participant drop-out and missing data, a challenge faced by most longitudinal studies. Also, whereas some of our assessment instruments are clinician-rated scales, the anxiety measurements (SAS and SIAS) are self-report scales, which may limit reliability. Furthermore, our analyses were limited by only capturing substance use in the month prior to each visit. To more accurately assess proximal relationships between substance use and clinical presentation, real-time data collected using methods like ecological momentary assessment are needed. Finally, although our finding of a temporal relationship between tobacco and cannabis use and anxiety is a step towards directionality, this relationship was still considered to be predictive precedence rather than causality (40). Future research should apply interventions targeted toward symptoms and measure their effects on substance use (or vice versa) to further our mechanistic understanding of the relationships between substance use, anxiety and depression, and psychosis-risk symptoms.

In conclusion, our findings shift the clinical focus towards a model where substance use in the CHR-P population is strongly linked with anxiety and level of social functioning. Biopsychosocial interventions should include strategies to alleviate anxiety symptoms, build skills for navigating social interactions that are pro-substance use, and incorporate baseline cannabis use into prognosis formulation and treatment planning.

## Data Availability

All data produced in the present study are available upon reasonable request to the authors.

## Acknowledgments

This work was supported by National Institutes of Health (NIH) grants U01 MH066134 to Dr. Addington, P50 MH066286 to Dr. Bearden, U01 MH081944 to Dr. Cadenhead, U01 MH081902 to Dr. Cannon, U01 MH081857 to Dr. Cornblatt, R01 MH076989 to Dr. Mathalon, U01 MH066069 to Dr. Perkins, U01 MH081928 to Dr. Stone, U01 MH081988 to Dr. Walker, U01 MH82022 to Dr. Woods, R01 MH116170 to Dr. Brady, and K23DA059690 to Dr. Ward.

## Competing Interests

The authors have no competing interests to disclose.

Note: Barbara Cornblatt and Larry Seidman passed away tragically before submission of this manuscript. Their colleagues wish to honor their contributions to the work posthumously.

