## Supplement tables for "Longitudinal Relationships Between Cannabis and Tobacco Use and Symptom Severity in Individuals at Clinical High Risk for Psychosis"

**Supplementary Methods**

**Participants**

*Inclusion and Exclusion Criteria*

Participants underwent clinical assessment at baseline and every 6 months for two years and upon conversion to psychosis (if applicable) from January 2009 to April 2013. Prior to participation, all participants provided written informed consent (or, if under age 18, informed assent with parental consent) in accordance with the institutional review boards of Beth Israel Deaconess Medical Center, Boston, Massachusetts; Emory University, Atlanta, Georgia; University of Calgary, Alberta, Canada; University of California, Los Angeles; University of California, San Diego; The University of North Carolina at Chapel Hill; Yale University, New Haven, Connecticut; and Zucker Hillside Hospital, New York.

CHR individuals were included if they met the Criteria for the Psychosis Risk Syndrome, based on the Structured Interview for Psychosis-Risk Syndromes (SIPS). If individuals were younger than 19 years, they were included based on criteria for schizotypal personality disorder or Criteria for the Psychosis Risk Syndrome. Individuals could meet any of four psychosis-risk syndrome criteria: 1) attenuated positive symptoms (APS), 2) brief intermittent psychotic symptoms (BIPS), 3) genetic risk and deterioration (GRD), or 4) youth and schizotypy criteria (YS). A CHR participant could meet criteria for multiple categories. The Structured Clinical Interview for DSM was used to exclude psychosis and to identify DSM-IV Axis I or cluster A personality disorders. Anyone with a lifetime Axis I psychotic disorder, estimated IQ less than 70 on both measures of IQ, a central nervous system disorder, or DSM-IV substance dependence in the past 6 months was excluded. Other nonpsychotic DSM-IV disorders were not exclusionary (e.g. depression, substance use disorders) unless they clearly caused or better explained prodromal symptoms. CHR individuals were permitted to take antipsychotic medication as long as they had not developed any psychotic symptoms prior to initiating medication. Healthy controls were not permitted to meet any prodromal criteria, have a history of a psychotic or cluster A personality disorder, or have a family history of a psychotic disorder in a first-degree relative. Prior to participation, all participants provided written informed consent (or, if under age 18, informed assent with parental consent) in accordance with institutional review boards.

**Measures**

*Clinical:* The Structured Clinical Interview for DSM was used to exclude psychosis and to identify DSM-IV Axis I or cluster A personality disorders. We categorized DSM-IV disorders into any affective disorders (major depressive disorder, dysthymic disorder, mood disorder not otherwise specified (NOS), or depressive disorder NOS) or anxiety disorders (panic disorder, posttraumatic stress disorder, obsessive-compulsive disorder, generalized anxiety disorder, anxiety disorder NOS, or phobias including simple phobias and social phobia).

*Psychiatric Symptoms*: Severity of psychosis symptoms at baseline was rated using the Scale of Psychosis-Risk Symptoms (SOPS). The SOPS is a clinician-administered assessment tool designed to evaluate early signs of psychosis in individuals at clinical high risk (CHR). Developed as part of the Structured Interview for Prodromal Syndromes (SIPS), the SOPS consists of four symptom domains: positive symptoms (e.g., unusual thought content, suspiciousness, perceptual abnormalities), negative symptoms (e.g., social withdrawal, decreased motivation), disorganized symptoms (e.g., trouble with communication and thinking), and general symptoms (e.g., anxiety, sleep disturbances). Anxiety was measured with the Self-Rating Anxiety Scale (SAS, “anxiety”) to assess self-report anxiety symptoms over the past few days and Social Interaction Anxiety Scale (SIAS, “social anxiety”) to assess self-report general social anxiety symptoms. Depressive symptoms were assessed using the Calgary Depression Scale for Schizophrenia (CDSS, “depression”), which is a clinician-administered tool designed to assess depressive symptoms specifically in individuals with schizophrenia.

*Substance Use*: Current substance use was measured using the Alcohol Use Scale/Drug Use Scale (AUS/DUS), which assesses substance use severity (abstinent, use without impairment, abuse, dependence, dependence with institutionalization) and frequency over the past 30 days for the following substances: tobacco, alcohol, marijuana, cocaine, opiates, phencyclidine (PCP), amphetamines, methylenedioxymethamphetamine (MDMA), gamma-hydroxybutarate, huffing, hallucinogens, and other substances. Substance use frequency was measured on an ordinal scale for tobacco in cigarettes per day (0 = no use, 1 = occasionally, 2 = less than 10 per day, 3 = 11-25 per day, 4 = more than 25 per day) and for all other substances (0 = no use, 1 = once or twice per month, 2 = 3-4 times per month, 3 = 1-2 times per week, 4 = 3-4 times per week, 5 = almost daily).

*Cognitive Performance*: Neurocognition was assessed with six domains from the MATRICS Consensus Cognitive Battery (MCCB): 1) Processing Speed (Trail Making Test Part A, Brief Assessment of Cognition in Schizophrenia, Symbol Coding); 2) Attention/Vigilance (Continuous Performance Test: Identical-Pairs); 3) Working Memory (Wechsler Memory Scale Spatial Span, Letter-number Span); 4) Verbal Learning (Hopkins Verbal Learning Test); 5) Visual Learning (Brief Visuospatial Memory Test; BVMT); 6) Reasoning and Problem-solving (Neuropsychological Assessment Battery: Mazes). The Mayer-Salovey-Caruso Emotional Intelligence Test: Managing Emotions (MSCEIT) was not included in NAPLS2. We calculated and z-scored overall performance and performance on each cognitive subdomain.

**Statistical Approach**

*Within-Visit Association:* We used linear mixed effect (LME) models with random intercept effect of subjects to assess the association between tobacco/cannabis use frequency and psychiatric symptoms, functioning, and cognition at each visit. Symptoms included total psychosis-risk and subscores (positive, negative, disorganization, general), anxiety, social anxiety, depression, social functioning, role functioning, global cognition and cognitive domain subscores. False discovery rate (FDR) adjusted p-values were computed separately across psychosis symptom subscores and across cognitive subdomain scores. Separate models were built for each pair of substance and symptom.

**Supplementary Results**

*Within-visit association*

Significant associations between tobacco or cannabis use frequency and clinical measurements within each visit were summarized in Figure S1. Within each visit, heavier current tobacco use was associated with worse psychosis-risk symptoms (β=0.971, p=0.0165), positive symptoms (β=0.398, p=0.0034, FDRp=0.0136), disorganization symptoms (β=0.216, p=0.0101, FDRp=0.024), general symptoms (p=0.0081, FDRp=0.024), and anxiety (β=1.315, p=0.0004), and with better social functioning (β=0.134, p=0.0013). Heavier cannabis use was associated with worse psychosis-risk symptoms (β=0.524, p=0.0325), positive symptoms (β=0.240, p=0.0035, FDRp=0.0140), general symptoms (β=0.198, p=0.0123, FDRp=0.0370), anxiety (β=0.577, p=0.0084), and depression (β=0.162, p=0.0378). No association was observed between tobacco or cannabis use frequency and cognitive measurements (Table S3). Post-hoc analysis did not find tobacco-by-cannabis interactive effect on current SOPS total, SOPS positive, SOPS general, or SAS (Table S3).

**Supplementary Figure 1**

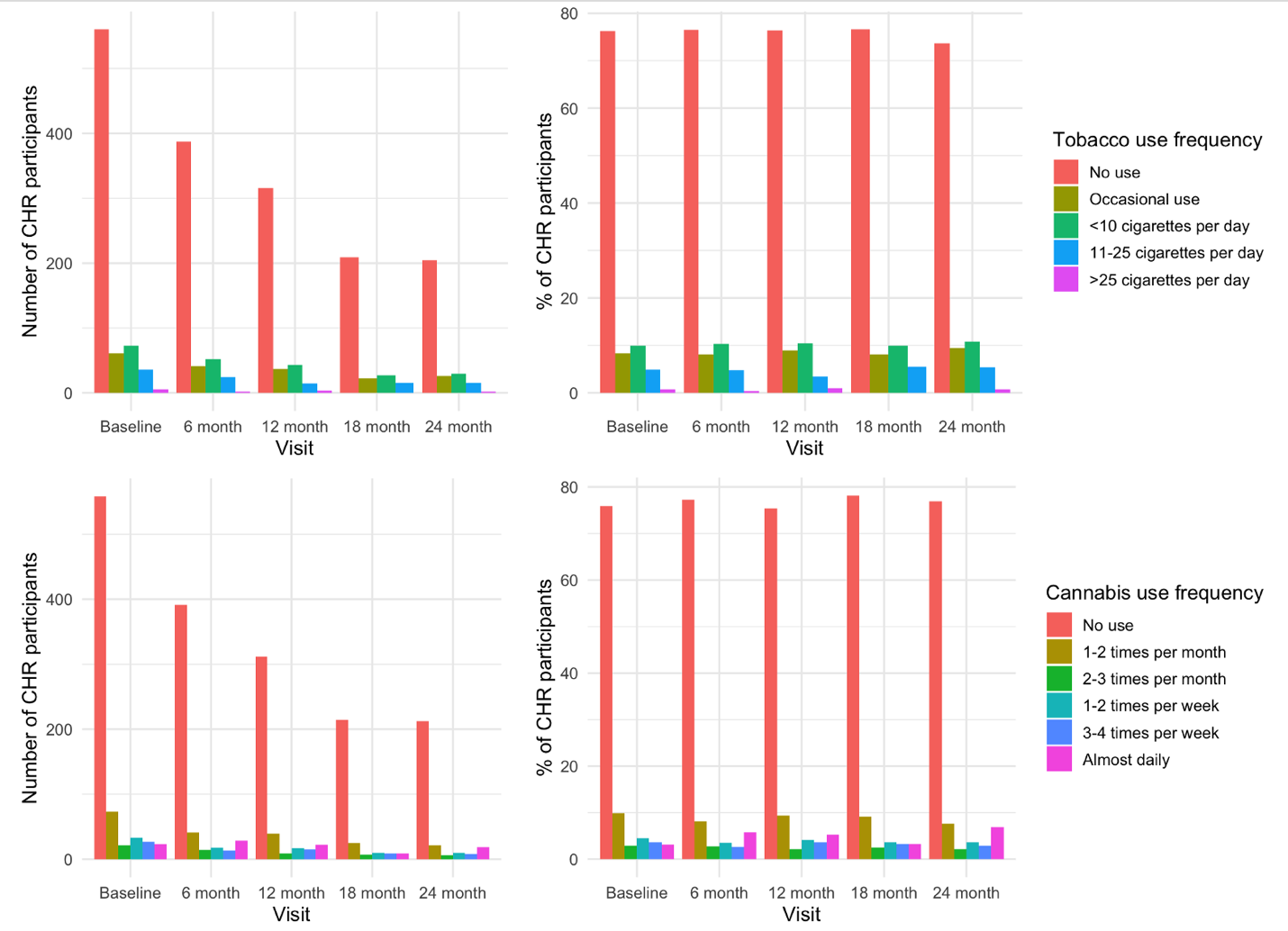

Supplementary Figure 1. Self-report tobacco and cannabis use frequent at each visit in NAPLS2. Both number of participants and percentage of participants among those with available substance use data were shown. Most CHR-P participants reported no use of tobacco or cannabis across all visits. CHR, clinical high risk for psychosis.

**Supplementary Figure 2**

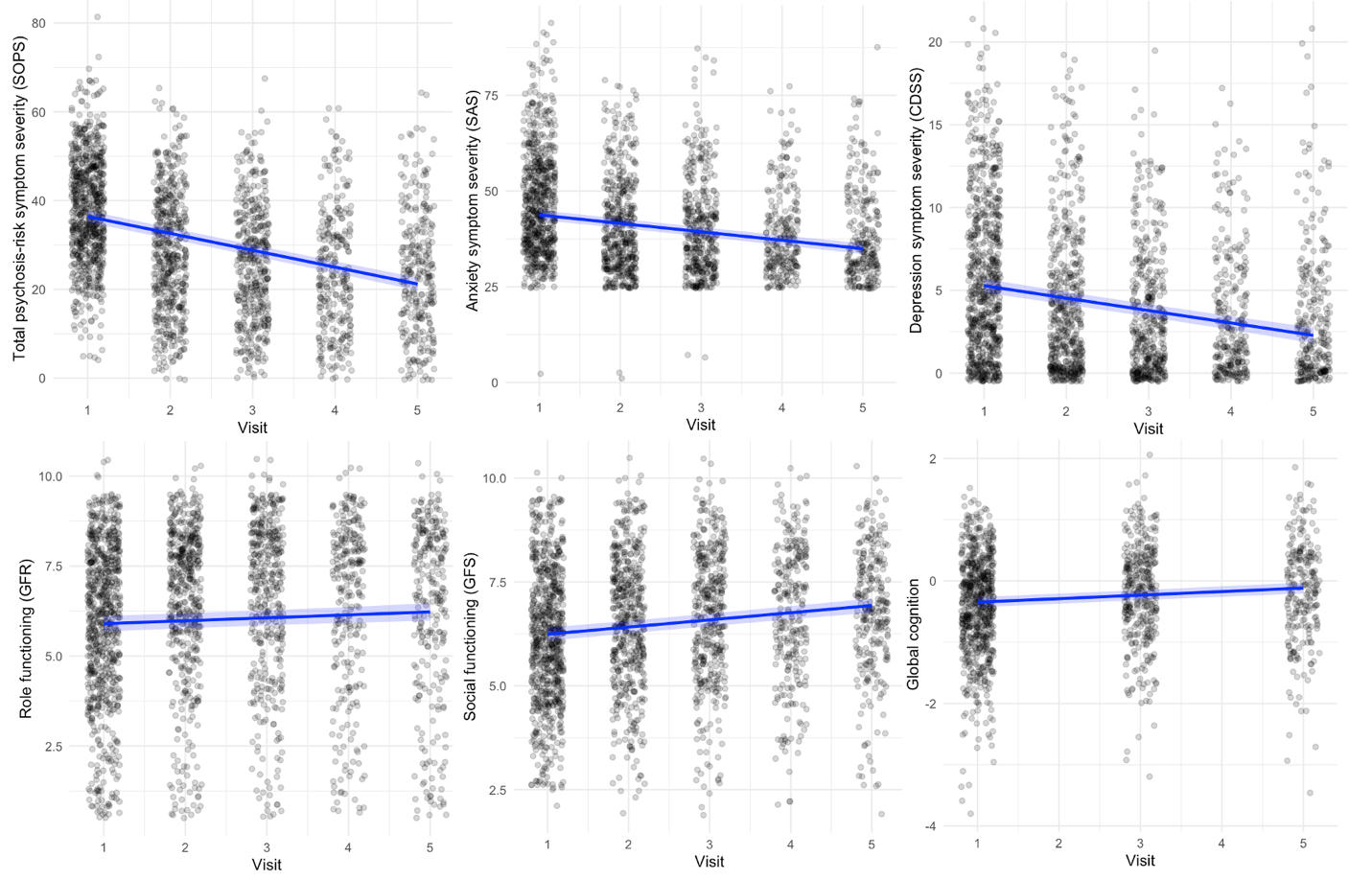

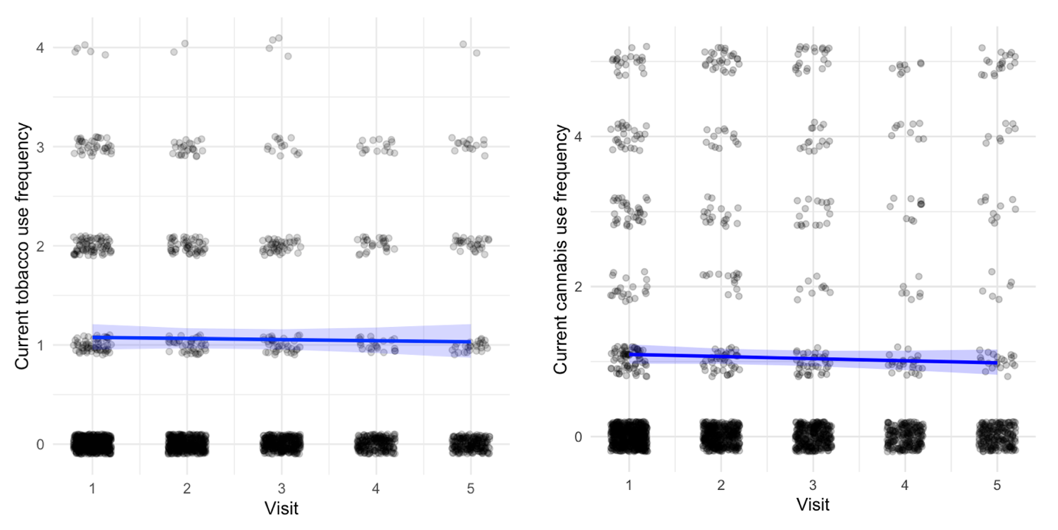

Supplementary Figure 2. Longitudinal trajectory of psychiatric symptoms (psychosis, anxiety, depression), functioning, global cognition, and substance use over the study period. Symptoms, functioning, and global cognition generally improved for CHR-P individuals over two years. Tobacco and cannabis use frequency did not change across all CHR-P individuals over two years. CDSS, Calgary Depression Scale for Schizophrenia; CHR-P, clinical high risk for psychosis; GFR, Global Functioning: Role Scale; GFS, Global Functioning: Social Scale; SAS, Self-Rating Anxiety Scale; SOPS, Scale of Psychosis-Risk Symptoms.

**Supplementary Figure 3**

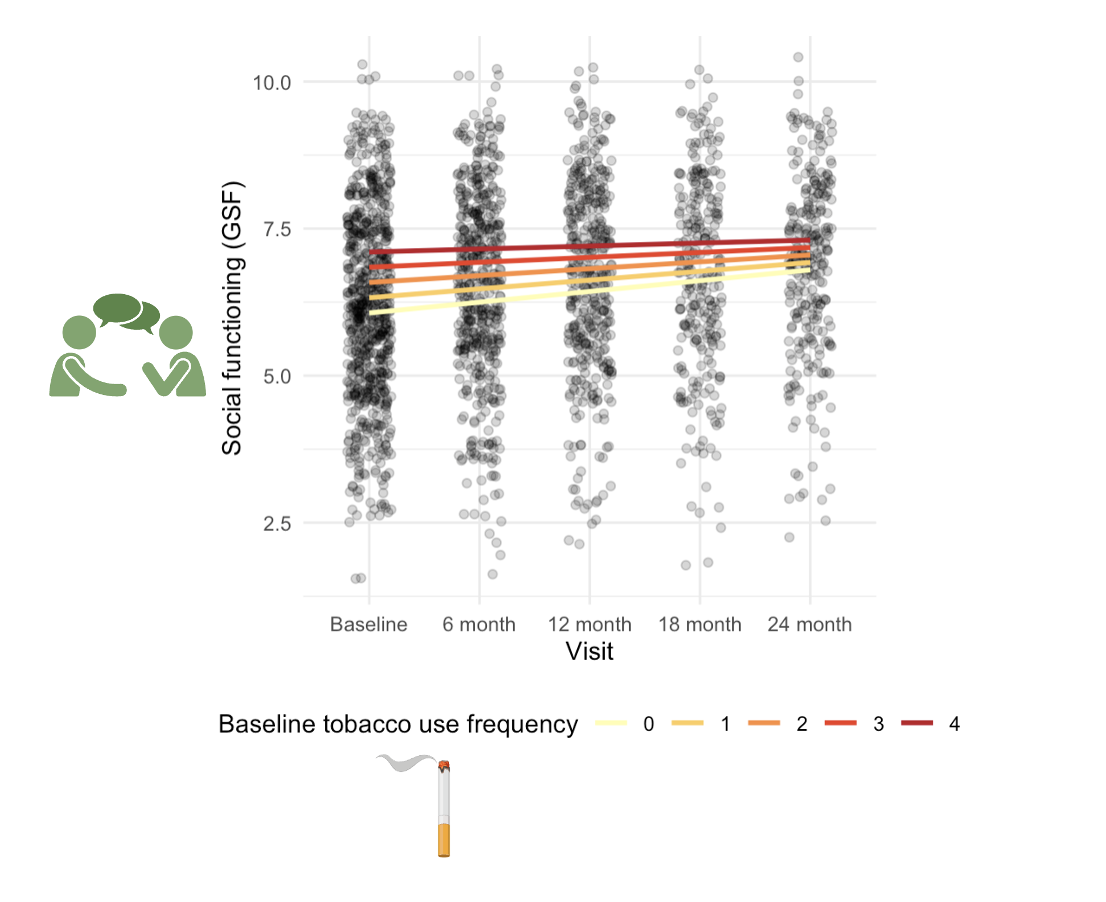

Supplementary Figure 3. Heavier baseline tobacco use predicts slower improvement of social functioning trajectory. GFS, Global Functioning: Social Scale.

**Supplementary Figure 4**

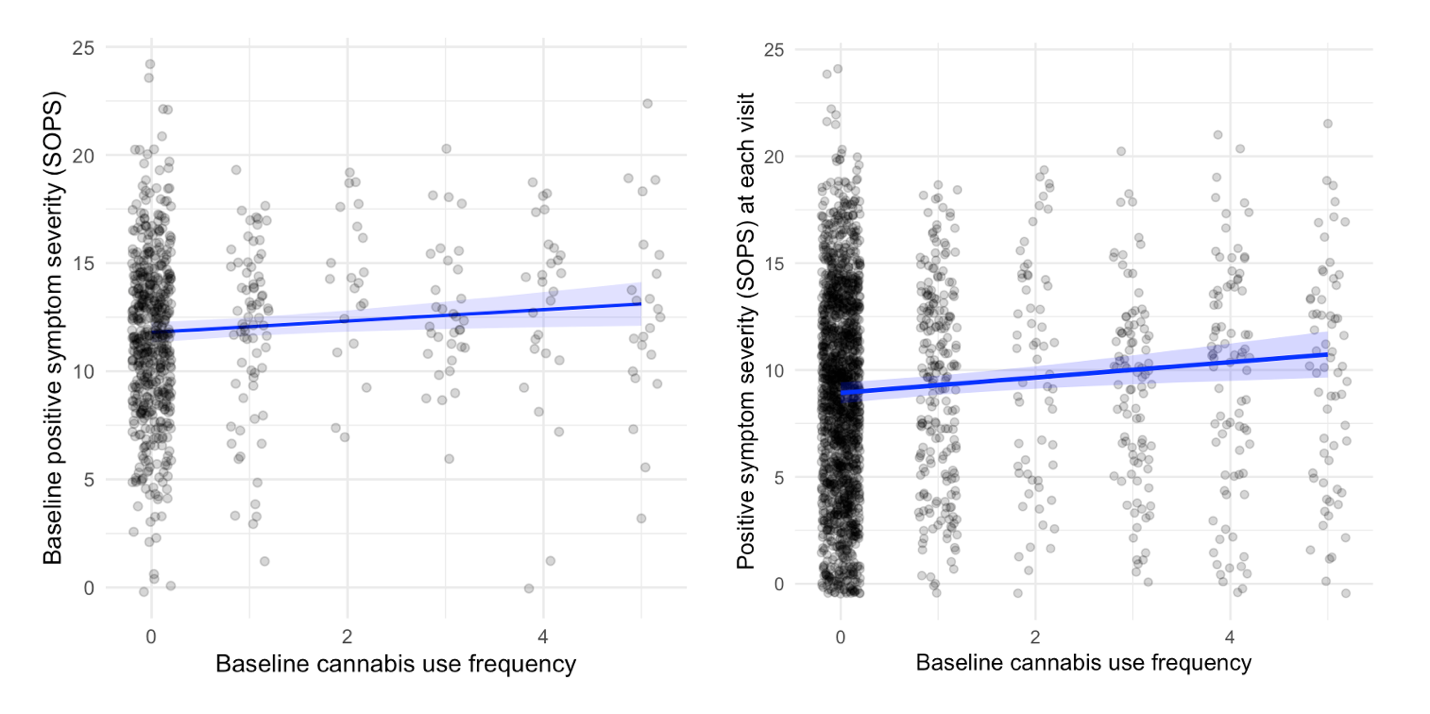

Supplementary Figure 4. Relationship between baseline cannabis use frequency and positive symptoms. Heavier cannabis use at baseline was associated with more severe positive symptoms both at baseline and at each visit over the study period.

**Supplementary Table 1: Data completion, substance use prevalence, and clinical measurements in CHR individuals in NAPLS-2**

| **Baseline demographics** | | | |  | | | | |
| --- | --- | --- | --- | --- | --- | --- | --- | --- |
|  | Age, mean (SD) | | | 18.96 (4.24) | | | | |
|  | Sex female, N (%) | | | 328 (42.93) | | | | |
|  | Taking antipsychotics, N (%) | | | 57 (7.46) | | | | |
|  | Antipsychotics dosage, mean (SD) | | | 211.14 (163.08) | | | | |
|  | Other SCID diagnosis, N (%) | | |  | | | | |
|  |  | *Any affective disorder* | | 294 (38.48) | | | | |
|  |  | Depression | | 252 (32.98) | | | | |
|  |  | Mania | | 42 (5.50) | | | | |
|  |  | Dysthymia | | 16 (2.09) | | | | |
|  |  | *Any anxiety disorder* | | 304 (39.79) | | | | |
|  |  | Social phobia | | 90 (11.78) | | | | |
|  |  | Panic disorder | | 76 (9.95) | | | | |
|  |  | Generalize anxiety disorder | | 70 (9.16) | | | | |
|  |  | Obsessive/compulsive disorder | | 43 (5.63) | | | | |
|  |  | *Schizotypal personality disorder* | | 104 (13.61) | | | | |
|  |  | *Missing data* | | 166 (21.73) | | | | |
| **Data completion and average symptom severity** | | | | | | | | |
|  | | | | Baseline | 6 month | 12 month | 18 month | 24 month |
| Current age, mean (SD) | | | | 18.96 (4.24) | 19.39 (4.29) | 20.17 (4.36) | 20.40 (4.41) | 21.32 (4.43) |
| Sex female, N (%) | | | | 328 (42.93) | 214 (42.41) | 174 (41.92) | 122 (44.36) | 123 (44.40) |
| AUS/DUS data completion, N | | | | 735 | 506 | 414 | 273 | 277 |
|  | Substance | | Use frequency, N (%) |  |  |  |  |  |
|  | Tobacco | | No use | 560 (76.19) | 387 (76.48) | 316 (76.32) | 209 (76.55) | 204 (73.64) |
|  |  | | Occasional use | 61 (8.30) | 41 (8.10) | 37 (8.93) | 22 (9.38) | 26 (9.38) |
|  |  | | <10 per day | 73 (9.93) | 52 (10.27) | 43 (10.38) | 27 (10.83) | 30 (10.83) |
|  |  | | 11-25 per day | 36 (4.90) | 24 (4.74) | 14 (3.38) | 15 (5.41) | 15 (5.41) |
|  |  | | > 25 per day | 5 (0.68) | 2 (0.39) | 4 (0.96) | 0 (0) | 2 (0.72) |
|  | Cannabis | | No use | 558 (75.91) | 391 (77.27) | 312 (75.36) | 214 (78.10) | 213 (76.90) |
|  |  | | 1-2 times per month | 73 (9.93) | 41 (8.10) | 39 (9.42) | 25 (9.12) | 21 (7.58) |
|  |  | | 3-4 times per month | 21 (2.85) | 14 (2.77) | 9 (2.17) | 7 (2.55) | 6 (2.17) |
|  |  | | 1-2 times per week | 33 (4.48) | 18 (3.56) | 17 (4.10) | 10 (3.65) | 10 (3.61) |
|  |  | | 3-4 times per week | 27 (3.67) | 13 (2.57) | 15 (3.62) | 9 (3.28) | 8 (2.89) |
|  |  | | Almost daily | 23 (3.13) | 29 (5.73) | 22 (5.31) | 9 (3.28) | 19 (6.86) |
| SOPS data completion, N | | | | 736 | 510 | 413 | 274 | 279 |
|  | Psychosis-risk symptoms, mean (SD) | | | 38.05 (12.15) | 28.41 (13.54) | 25.57 (13.05) | 23.66 (13.78) | 23.56 (13.79) |
|  | Positive symptoms, mean (SD) | | | 11.91 (3.82) | 8.57 (4.50) | 7.75 (4.61) | 6.76 (4.60) | 6.63 (4.61) |
|  | Negative symptoms, mean (SD) | | | 11.89 (6.07) | 9.16 (5.91) | 8.31 (5.80) | 8.06 (6.10) | 7.85 (5.84) |
|  | Disorganization symptoms, mean (SD) | | | 5.16 (3.16) | 3.99 (2.94) | 3.53 (2.80) | 3.12 (2.59) | 3.35 (2.81) |
|  | General symptoms, mean (SD) | | | 9.16 (4.27) | 6.70 (4.47) | 5.98 (4.17) | 5.77 (4.41) | 5.73 (4.38) |
| SAS data completion, N | | | | 694 | 485 | 407 | 255 | 261 |
|  | Anxiety (SAS), mean (SD) | | | 47.37 (13.64) | 41.08 (12.55) | 40.32 (12.60) | 38.92 (11.09) | 38.68 (12.05) |
| SIAS data completion, N | | | | 690 | 487 | 406 | 253 | 259 |
|  | Social anxiety (SIAS), mean (SD) | | | 30.80 (17.41) | 26.53 (16.79) | 25.12 (17.52) | 25.94 (17.09) | 24.07 (17.00) |
| CDSS data completion, N | | | | 721 | 503 | 413 | 274 | 277 |
|  | Depression (CDSS) total, mean (SD) | | | 5.82 (4.76) | 4.23 (4.34) | 3.55 (3.80) | 3.51 (3.79) | 3.42 (4.06) |
| GFR/GFS data completion, N | | | | 753 | 510 | 413 | 274 | 276 |
|  | Role functioning (GFR), mean (SD) | | | 5.95 (2.15) | 6.53 (2.11) | 6.61 (2.17) | 6.45 (2.35) | 6.36 (2.44) |
|  | Social functioning (GFS), mean (SD) | | | 6.18 (1.57) | 6.58 (1.57) | 6.69 (1.58) | 6.74 (1.59) | 6.85 (1.49) |
| MATRICS data completion, N | | | | 687 |  | 360 |  | 235 |
|  | Global cognition, mean (SD) | | | -0.42 (0.78) |  | -0.18 (0.74) |  | -0.16 (0.79) |
|  | Processing speed, mean (SD) | | | -0.59 (1.12) |  | -0.27 (1.13) |  | -0.24 (1.19) |
|  | Working memory, mean (SD) | | | -0.52 (1.13) |  | -0.27 (1.10) |  | -0.29 (1.12) |
|  | Attention, mean (SD) | | | -0.57 (1.08) |  | -0.20 (1.04) |  | -0.14 (1.22) |
|  | Verbal learning, mean (SD) | | | -0.38 (1.30) |  | -0.15 (1.17) |  | -0.07 (1.16) |
|  | Visual learning, mean (SD) | | | -0.38 (1.15) |  | -0.25 (1.11) |  | -0.25 (1.05) |
|  | Reasoning and problem solving, mean (SD) | | | -0.18 (1.11) |  | 0.05 (1.03) |  | -0.02 (1.07) |

**Supplementary Table 2: Longitudinal trajectory**

| Response | Fixed effect | | | | | | Random effect | |
| --- | --- | --- | --- | --- | --- | --- | --- | --- |
|  | Term | Estimate | SE | t-value | Chi-square | p-value | Term | Variance (SD) |
| SOPS total | Visit | -3.804 | 0.1607 | -23.670 | 560.260 | <0.0001 | Subject | 96.78 (9.838) |
|  | Intercept | 35.803 | 1.884 | 19.000 |  |  | Residual | 76.93 (8.771) |
| SAS | Visit | -2.2043 | 0.1542 | -14.292 | 204.2735 | <0.0001 | Subject | 92.01 (9.592) |
|  | Intercept | 41.0220 | 1.8094 | 22.671 |  |  | Residual | 65.93 (8.120) |
| CDSS | Visit | -0.7498 | 0.0579 | -12.942 | 167.4944 | <0.0001 | Subject | 7.336 (2.709) |
|  | Intercept | 3.42892 | 0.6058 | 5.660 |  |  | Residual | 10.784 (3.284) |
| GFR | Visit | 0.0816 | 0.0268 | 3.043 | 9.261 | 0.00234 | Subject | 2.686 (1.639) |
|  | Intercept | 6.098 | 0.311 | 19.625 |  |  | Residual | 2.147 (1.465) |
| GFS | Visit | 0.173 | 0.0171 | 10.052 | 101.016 | <0.0001 | Subject | 1.630 (1.277) |
|  | Intercept | 5.974 | 0.217 | 27.521 |  |  | Residual | 0.823 (0.907) |
| Global cognition | Visit | 0.0578 | 0.00762 | 7.591 | 57.622 | <0.0001 | Subject | 0.499 (0.707) |
|  | Intercept | -0.740 | 0.105 | -7.060 |  |  | Residual | 0.0948 (0.308) |

| Response | Term | Coefficient | SE | Wald z-score | p-value |
| --- | --- | --- | --- | --- | --- |
| Tobacco | Visit | -0.0146 | 0.0343 | -0.43 | 0.671 |
| Cannabis | Visit | -0.0368 | 0.0354 | -1.04 | 0.2982 |

**Supplementary Table 3: single visit association**

Tobacco

| Response | Fixed effect | | | | | | Random effect | | |
| --- | --- | --- | --- | --- | --- | --- | --- | --- | --- |
|  | Term | Estimate | SE | t-value | p-value | FDRp | Term | Variance | SD |
| SOPS total | Intercept | 39.822 | 2.080 | 19.145 |  |  | Subject | 114.241 | 10.688 |
|  | Tobacco | 0.971 | 0.405 | 2.397 | **0.0165** |  | Residual | 98.318 | 9.916 |
| SOPS positive | Intercept | 12.640 | 0.685 | 18.461 |  |  | Subject | 11.843 | 3.441 |
|  | Tobacco | 0.398 | 0.136 | 2.930 | **0.0034** | **0.0136** | Residual | 11.648 | 3.413 |
| SOPS negative | Intercept | 13.173 | 0.885 | 14.889 |  |  | Subject | 23.106 | 4.807 |
|  | Tobacco | 0.036 | 0.171 | 0.209 | 0.835 | 0.835 | Residual | 16.168 | 4.021 |
| SOPS disorganization | Intercept | 5.791 | 0.437 | 13.243 |  |  | Subject | 5.785 | 2.405 |
|  | Tobacco | 0.216 | 0.084 | 2.571 | **0.0101** | **0.0244** | Residual | 3.858 | 1.964 |
| SOPS general | Intercept | 8.091 | 0.651 | 12.424 |  |  | Subject | 9.511 | 3.084 |
|  | Tobacco | 0.341 | 0.129 | 2.647 | **0.0081** | **0.0244** | Residual | 11.215 | 3.349 |
| SAS | Intercept | 43.675 | 1.867 | 23.396 |  |  | Subject | 95.876 | 9.792 |
|  | Tobacco | 1.315 | 0.369 | 3.564 | **0.0004** |  | Residual | 73.210 | 8.556 |
| SIAS | Intercept | 25.943 | 2.445 | 10.612 |  |  | Subject | 196.238 | 14.008 |
|  | Tobacco | -0.447 | 0.466 | -0.960 | 0.3372 |  | Residual | 103.659 | 10.181 |
| CDSS | Intercept | 3.654 | 0.624 | 5.856 |  |  | Subject | 7.520 | 2.742 |
|  | Tobacco | 0.151 | 0.125 | 1.204 | 0.2286 |  | Residual | 11.774 | 3.431 |
| GFR | Intercept | 6.002 | 0.310 | 19.391 |  |  | Subject | 2.666 | 1.633 |
|  | Tobacco | -0.119 | 0.061 | -1.959 | 0.0501 |  | Residual | 2.164 | 1.471 |
| GFS | Intercept | 5.684 | 0.218 | 26.121 |  |  | Subject | 1.625 | 1.275 |
|  | Tobacco | 0.134 | 0.042 | 3.222 | **0.0013** |  | Residual | 0.871 | 0.933 |
| Global cognition | Intercept | -0.906 | 0.105 | -8.660 |  |  | Subject | 0.511 | 0.715 |
|  | Tobacco | -0.021 | 0.021 | -0.998 | 0.318 |  | Residual | 0.101 | 0.318 |
| Processing speed | Intercept | -1.076 | 0.168 | -6.406 |  |  | Subject | 0.873 | 0.935 |
|  | Tobacco | -0.003 | 0.036 | -0.081 | 0.935 | 1 | Residual | 0.400 | 0.632 |
| Working memory | Intercept | -1.038 | 0.164 | -6.335 |  |  | Subject | 0.946 | 0.973 |
|  | Tobacco | -0.018 | 0.035 | -0.503 | 0.615 | 1 | Residual | 0.332 | 0.576 |
| Attention | Intercept | -1.116 | 0.167 | -6.663 |  |  | Subject | 0.826 | 0.909 |
|  | Tobacco | -0.046 | 0.037 | -1.248 | 0.212 | 1 | Residual | 0.397 | 0.630 |
| Verbal learning | Intercept | -0.614 | 0.192 | -3.202 |  |  | Subject | 0.919 | 0.959 |
|  | Tobacco | -0.064 | 0.042 | -1.525 | 0.127 | 0.763 | Residual | 0.646 | 0.804 |
| Visual learning | Intercept | -0.583 | 0.171 | -3.407 |  |  | Subject | 0.711 | 0.843 |
|  | Tobacco | -0.020 | 0.037 | -0.522 | 0.602 | 1 | Residual | 0.527 | 0.726 |
| Problem reasoning | Intercept | -0.272 | 0.160 | -1.694 |  |  | Subject | 0.756 | 0.870 |
|  | Tobacco | -0.014 | 0.035 | -0.405 | 0.685 | 1 | Residual | 0.384 | 0.620 |

Cannabis

| Response | Fixed effect | | | | | | Random effect | | |
| --- | --- | --- | --- | --- | --- | --- | --- | --- | --- |
|  | Term | Estimate | SE | t-value | p-value | FDRp | Term | Variance | SD |
| SOPS total | Intercept | 39.694 | 2.079 | 19.093 |  |  | Subject | 114.241 | 10.688 |
|  | Cannabis | 0.524 | 0.245 | 2.139 | **0.0325** |  | Residual | 98.318 | 9.916 |
| SOPS positive | Intercept | 12.551 | 0.681 | 18.420 |  |  | Subject | 11.843 | 3.441 |
|  | Cannabis | 0.240 | 0.082 | 2.920 | **0.0035** | **0.0140** | Residual | 11.648 | 3.413 |
| SOPS negative | Intercept | 13.170 | 0.884 | 14.904 |  |  | Subject | 23.106 | 4.807 |
|  | Cannabis | 0.012 | 0.102 | 0.119 | 0.9053 | 0.9053 | Residual | 16.168 | 4.021 |
| SOPS disorganization | Intercept | 5.750 | 0.437 | 13.161 |  |  | Subject | 5.785 | 2.405 |
|  | Cannabis | 0.081 | 0.050 | 1.620 | 0.1051 | 0.2103 | Residual | 3.858 | 1.964 |
| SOPS general | Intercept | 8.071 | 0.652 | 12.379 |  |  | Subject | 9.511 | 3.084 |
|  | Cannabis | 0.198 | 0.079 | 2.502 | **0.0123** | **0.0370** | Residual | 11.215 | 3.349 |
| SAS | Intercept | 43.565 | 1.871 | 23.279 |  |  | Subject | 95.876 | 9.792 |
|  | Cannabis | 0.577 | 0.219 | 2.635 | **0.0084** |  | Residual | 73.210 | 8.556 |
| SIAS | Intercept | 26.019 | 2.445 | 10.642 |  |  | Subject | 196.238 | 14.008 |
|  | Cannabis | -0.123 | 0.273 | -0.450 | 0.6526 |  | Residual | 103.659 | 10.181 |
| CDSS | Intercept | 3.643 | 0.623 | 5.849 |  |  | Subject | 7.520 | 2.742 |
|  | Cannabis | 0.162 | 0.078 | 2.077 | **0.0378** |  | Residual | 11.774 | 3.431 |
| GFR | Intercept | 6.010 | 0.310 | 19.387 |  |  | Subject | 2.666 | 1.633 |
|  | Cannabis | -0.028 | 0.037 | -0.772 | 0.4402 |  | Residual | 2.164 | 1.471 |
| GFS | Intercept | 5.668 | 0.218 | 25.979 |  |  | Subject | 1.625 | 1.275 |
|  | Cannabis | 0.035 | 0.024 | 1.444 | 0.1487 |  | Residual | 0.871 | 0.933 |
| Global cognition | Intercept | -0.903 | 0.104 | -8.648 |  |  | Subject | 0.511 | 0.715 |
|  | Cannabis | 0.015 | 0.013 | 1.169 | 0.2424 |  | Residual | 0.101 | 0.318 |
| Processing speed | Intercept | -1.074 | 0.168 | -6.407 |  |  | Subject | 0.873 | 0.935 |
|  | Cannabis | 0.026 | 0.023 | 1.160 | 0.2460 | 0.8506 | Residual | 0.400 | 0.632 |
| Working memory | Intercept | -1.031 | 0.163 | -6.318 |  |  | Subject | 0.946 | 0.973 |
|  | Cannabis | 0.028 | 0.021 | 1.329 | 0.1838 | 0.8506 | Residual | 0.332 | 0.576 |
| Attention | Intercept | -1.112 | 0.168 | -6.637 |  |  | Subject | 0.826 | 0.909 |
|  | Cannabis | 0.031 | 0.023 | 1.372 | 0.1701 | 0.8506 | Residual | 0.397 | 0.630 |
| Verbal learning | Intercept | -0.612 | 0.192 | -3.188 |  |  | Subject | 0.919 | 0.959 |
|  | Cannabis | -0.010 | 0.027 | -0.364 | 0.7161 | 0.9645 | Residual | 0.646 | 0.804 |
| Visual learning | Intercept | -0.581 | 0.171 | -3.401 |  |  | Subject | 0.711 | 0.843 |
|  | Cannabis | 0.037 | 0.024 | 1.552 | 0.1206 | 0.7233 | Residual | 0.527 | 0.726 |
| Problem reasoning | Intercept | -0.266 | 0.160 | -1.658 |  |  | Subject | 0.756 | 0.870 |
|  | Cannabis | 0.015 | 0.022 | 0.703 | 0.4823 | 0.9645 | Residual | 0.384 | 0.620 |

**Supplementary Table 4: current substance use predict future clinical measurements**

Tobacco

| Response | Fixed effect | | | | | | Random effect | | |
| --- | --- | --- | --- | --- | --- | --- | --- | --- | --- |
|  | Term | Estimate | SE | t-value | p-value | FDRp | Term | Variance | SD |
| Future SOPS total | Intercept | 8.098 | 1.901 | 4.259 |  |  | Subject | 31.459 | 5.609 |
|  | Tobacco | 0.032 | 0.381 | 0.085 | 0.932 |  | Residual | 75.208 | 8.672 |
| Future SOPS positive | Intercept | 2.775 | 0.661 | 4.195 |  |  | Subject | 3.965 | 1.991 |
|  | Tobacco | -0.034 | 0.134 | -0.255 | 0.798 | 0.963 | Residual | 9.253 | 3.042 |
| Future SOPS negative | Intercept | 1.578 | 0.698 | 2.262 |  |  | Subject | 1.548 | 1.244 |
|  | Tobacco | -0.172 | 0.147 | -1.173 | 0.240 | 0.963 | Residual | 16.759 | 4.094 |
| Future SOPS disorganization | Intercept | 0.743 | 0.343 | 2.166 |  |  | Subject | 0.303 | 0.551 |
|  | Tobacco | 0.067 | 0.072 | 0.928 | 0.353 | 0.963 | Residual | 4.203 | 2.050 |
| Future SOPS general | Intercept | 1.439 | 0.615 | 2.338 |  |  | Subject | 2.018 | 1.420 |
|  | Tobacco | 0.150 | 0.129 | 1.161 | 0.245 | 0.963 | Residual | 11.203 | 3.347 |
| Future SAS | Intercept | 14.838 | 1.714 | 8.659 |  |  | Subject | 4.004 | 2.001 |
|  | Tobacco | 0.712 | 0.334 | 2.134 | **0.033** |  | Residual | 85.274 | 9.234 |
| Future SIAS | Intercept | 0.474 | 1.876 | 0.253 |  |  | Subject | 0.000 | 0.000 |
|  | Tobacco | -0.333 | 0.402 | -0.829 | 0.407 |  | Residual | 139.712 | 11.820 |
| Future CDSS | Intercept | 0.566 | 0.612 | 0.925 |  |  | Subject | 3.256 | 1.804 |
|  | Tobacco | -0.004 | 0.132 | -0.030 | 0.975 |  | Residual | 9.541 | 3.089 |
| Future GFR | Intercept | 2.706 | 0.288 | 9.386 |  |  | Subject | 0.000 | 0.000 |
|  | Tobacco | 0.012 | 0.056 | 0.217 | 0.828 |  | Residual | 2.940 | 1.715 |
| Future GFS | Intercept | 2.138 | 0.204 | 10.462 |  |  | Subject | 0.000 | 0.000 |
|  | Tobacco | 0.055 | 0.035 | 1.558 | 0.119 |  | Residual | 1.148 | 1.071 |
| Future global cognition | Intercept | 0.209 | 0.086 | 2.414 |  |  | Subject | 0.000 | 0.000 |
|  | Tobacco | -0.005 | 0.020 | -0.247 | 0.805 |  | Residual | 0.158 | 0.398 |
| Future processing speed | Intercept | 0.091 | 0.170 | 0.534 |  |  | Subject | 0.000 | 0.000 |
|  | Tobacco | 0.036 | 0.039 | 0.941 | 0.346 | 1.00 | Residual | 0.616 | 0.785 |
| Future working memory | Intercept | 0.467 | 0.159 | 2.932 |  |  | Subject | 0.000 | 0.000 |
|  | Tobacco | -0.024 | 0.036 | -0.662 | 0.507 | 1.00 | Residual | 0.541 | 0.735 |
| Future attention | Intercept | 0.341 | 0.180 | 1.897 |  |  | Subject | 0.000 | 0.000 |
|  | Tobacco | -0.005 | 0.040 | -0.116 | 0.907 | 1.00 | Residual | 0.634 | 0.796 |
| Future verbal learning | Intercept | -0.456 | 0.207 | -2.198 |  |  | Subject | 0.000 | 0.000 |
|  | Tobacco | -0.031 | 0.047 | -0.671 | 0.501 | 1.00 | Residual | 0.913 | 0.955 |
| Future visual learning | Intercept | -0.175 | 0.195 | -0.898 |  |  | Subject | 0.000 | 0.000 |
|  | Tobacco | -0.019 | 0.043 | -0.440 | 0.659 | 1.00 | Residual | 0.772 | 0.879 |
| Future problem reasoning | Intercept | 0.510 | 0.161 | 3.167 |  |  | Subject | 0.000 | 0.000 |
|  | Tobacco | 0.021 | 0.036 | 0.590 | 0.555 | 1.00 | Residual | 0.542 | 0.736 |

Cannabis

| Response | Fixed effect | | | | | | Random effect | | |
| --- | --- | --- | --- | --- | --- | --- | --- | --- | --- |
|  | Term | Estimate | SE | t-value | p-value | FDRp | Term | Variance | SD |
| Future SOPS total | Intercept | 7.916 | 1.892 | 4.184 |  |  | Subject | 30.607 | 5.532 |
|  | Cannabis | 0.079 | 0.244 | 0.325 | 0.744 |  | Residual | 75.665 | 8.699 |
| Future SOPS positive | Intercept | 2.720 | 0.659 | 4.128 |  |  | Subject | 3.872 | 1.968 |
|  | Cannabis | 0.032 | 0.086 | 0.368 | 0.712 | 1.00 | Residual | 9.309 | 3.051 |
| Future SOPS negative | Intercept | 1.609 | 0.700 | 2.297 |  |  | Subject | 1.663 | 1.289 |
|  | Cannabis | -0.053 | 0.097 | -0.543 | 0.587 | 1.00 | Residual | 16.678 | 4.084 |
| Future SOPS disorganization | Intercept | 0.719 | 0.341 | 2.105 |  |  | Subject | 0.288 | 0.536 |
|  | Cannabis | 0.060 | 0.048 | 1.258 | 0.208 | 0.834 | Residual | 4.209 | 2.051 |
| Future SOPS general | Intercept | 1.379 | 0.613 | 2.251 |  |  | Subject | 1.943 | 1.394 |
|  | Cannabis | 0.058 | 0.084 | 0.691 | 0.489 | 1.00 | Residual | 11.265 | 3.356 |
| Future SAS | Intercept | 14.314 | 1.694 | 8.450 |  |  | Subject | 2.827 | 1.681 |
|  | Cannabis | 0.511 | 0.217 | 2.356 | **0.018** |  | Residual | 86.313 | 9.290 |
| Future SIAS | Intercept | 0.507 | 1.876 | 0.270 |  |  | Subject | 0.000 | 0.000 |
|  | Cannabis | -0.009 | 0.264 | -0.034 | 0.972 |  | Residual | 139.676 | 11.818 |
| Future CDSS | Intercept | 0.533 | 0.608 | 0.876 |  |  | Subject | 3.098 | 1.760 |
|  | Cannabis | 0.129 | 0.085 | 1.514 | 0.129 |  | Residual | 9.629 | 3.103 |
| Future GFR | Intercept | 2.725 | 0.289 | 9.443 |  |  | Subject | 0.000 | 0.000 |
|  | Cannabis | -0.049 | 0.037 | -1.303 | 0.192 |  | Residual | 2.945 | 1.716 |
| Future GFS | Intercept | 2.117 | 0.203 | 10.452 |  |  | Subject | 0.000 | 0.000 |
|  | Cannabis | 0.060 | 0.023 | 2.602 | **0.009** |  | Residual | 1.142 | 1.069 |
| Future global cognition | Intercept | 0.209 | 0.086 | 2.428 |  |  | Subject | 0.000 | 0.000 |
|  | Cannabis | -0.004 | 0.013 | -0.286 | 0.775 |  | Residual | 0.158 | 0.397 |
| Future processing speed | Intercept | 0.084 | 0.170 | 0.497 |  |  | Subject | 0.000 | 0.000 |
|  | Cannabis | 0.031 | 0.026 | 1.182 | 0.237 | 0.948 | Residual | 0.615 | 0.784 |
| Future working memory | Intercept | 0.470 | 0.159 | 2.954 |  |  | Subject | 0.000 | 0.000 |
|  | Cannabis | -0.004 | 0.024 | -0.166 | 0.868 | 1.00 | Residual | 0.540 | 0.735 |
| Future attention | Intercept | 0.339 | 0.180 | 1.887 |  |  | Subject | 0.000 | 0.000 |
|  | Cannabis | 0.010 | 0.027 | 0.386 | 0.699 | 1.00 | Residual | 0.633 | 0.796 |
| Future verbal learning | Intercept | -0.450 | 0.207 | -2.173 |  |  | Subject | 0.000 | 0.000 |
|  | Cannabis | 0.007 | 0.032 | 0.234 | 0.815 | 1.00 | Residual | 0.912 | 0.955 |
| Future visual learning | Intercept | -0.171 | 0.194 | -0.880 |  |  | Subject | 0.000 | 0.000 |
|  | Cannabis | -0.049 | 0.029 | -1.700 | 0.089 | 0.534 | Residual | 0.767 | 0.876 |
| Future problem reasoning | Intercept | 0.505 | 0.160 | 3.150 |  |  | Subject | 0.000 | 0.000 |
|  | Cannabis | 0.040 | 0.024 | 1.668 | 0.095 | 0.534 | Residual | 0.539 | 0.734 |

**Supplementary Table 5: current clinical measurements predict future substance use**

| Response | Term | Coefficient | SE | Wald z-score | p-value | FDRp |
| --- | --- | --- | --- | --- | --- | --- |
| Future tobacco | SOPS total | -0.004 | 0.005 | -0.815 | 0.415 |  |
|  | SOPS positive | -0.003 | 0.016 | -0.215 | 0.830 | 0.891 |
|  | SOPS negative | -0.026 | 0.013 | -1.961 | 0.0498 | 0.199 |
|  | SOPS disorganization | -0.025 | 0.024 | -1.020 | 0.308 | 0.891 |
|  | SOPS general | 0.018 | 0.017 | 1.043 | 0.297 | 0.891 |
|  | SAS | 0.010 | 0.005 | 1.831 | 0.067 |  |
|  | SIAS | -0.004 | 0.005 | -0.844 | 0.399 |  |
|  | CDSS | -0.005 | 0.017 | -0.290 | 0.772 |  |
|  | GFR | -0.011 | 0.032 | -0.360 | 0.719 |  |
|  | GFS | 0.178 | 0.051 | 3.502 | **<0.001** |  |
|  | Global cognition | 0.074 | 0.149 | 0.494 | 0.621 |  |
|  | Processing speed | -0.087 | 0.089 | -0.979 | 0.328 | 1.00 |
|  | Working memory | 0.104 | 0.090 | 1.154 | 0.249 | 1.00 |
|  | Attention | -0.033 | 0.091 | -0.365 | 0.715 | 1.00 |
|  | Verbal learning | 0.112 | 0.097 | 1.152 | 0.249 | 1.00 |
|  | Visual learning | 0.118 | 0.104 | 1.135 | 0.256 | 1.00 |
|  | Problem reasoning | -0.057 | 0.104 | -0.545 | 0.586 | 1.00 |
| Future cannabis | SOPS total | 0.004 | 0.005 | 0.824 | 0.410 |  |
|  | SOPS positive | 0.028 | 0.017 | 1.619 | 0.105 | 0.422 |
|  | SOPS negative | -0.009 | 0.011 | -0.815 | 0.415 | 0.675 |
|  | SOPS disorganization | 0.022 | 0.023 | 0.959 | 0.337 | 0.675 |
|  | SOPS general | 0.020 | 0.015 | 1.296 | 0.195 | 0.584 |
|  | SAS | 0.009 | 0.006 | 1.555 | 0.120 |  |
|  | SIAS | -0.003 | 0.004 | -0.701 | 0.483 |  |
|  | CDSS | 0.016 | 0.015 | 1.051 | 0.293 |  |
|  | GFR | -0.020 | 0.033 | -0.622 | 0.534 |  |
|  | GFS | 0.162 | 0.047 | 3.447 | **<0.001** |  |
|  | Global cognition | 0.166 | 0.147 | 1.135 | 0.256 |  |
|  | Processing speed | 0.131 | 0.091 | 1.441 | 0.150 | 0.898 |
|  | Working memory | 0.099 | 0.097 | 1.017 | 0.309 | 0.928 |
|  | Attention | -0.062 | 0.106 | -0.589 | 0.556 | 1.000 |
|  | Verbal learning | 0.010 | 0.083 | 0.121 | 0.904 | 1.000 |
|  | Visual learning | 0.132 | 0.102 | 1.298 | 0.194 | 0.917 |
|  | Problem reasoning | 0.137 | 0.103 | 1.330 | 0.183 | 0.917 |

**Supplementary Table 6: Baseline substance use modify trajectory of clinical measurements**

Tobacco

| Response | Fixed effect |  |  |  |  |  | Random effect |  |  |
| --- | --- | --- | --- | --- | --- | --- | --- | --- | --- |
|  | Term | Estimate | SE | t-value | p-value | FDRp | Term | Variance | SD |
| SOPS total | Intercept | 39.135 | 0.716 | 54.645 |  |  | Subject | 98.287 | 9.914 |
|  | Baseline tobacco x visit | 0.105 | 0.160 | 0.658 | 0.510 | 1.00 | Residual | 76.665 | 8.756 |
| SOPS positive | Intercept | 12.373 | 0.235 | 52.752 |  |  | Subject | 10.218 | 3.197 |
|  | Baseline tobacco x visit | 0.057 | 0.054 | 1.062 | 0.288 | 1.00 | Residual | 8.841 | 2.973 |
| SOPS negative | Intercept | 12.928 | 0.325 | 39.757 |  |  | Subject | 21.656 | 4.654 |
|  | Baseline tobacco x visit | 0.076 | 0.070 | 1.093 | 0.274 | 1.00 | Residual | 14.398 | 3.794 |
| SOPS disorganization | Intercept | 5.280 | 0.162 | 32.647 |  |  | Subject | 5.356 | 2.314 |
|  | Baseline tobacco x visit | 0.018 | 0.035 | 0.529 | 0.597 | 1.00 | Residual | 3.567 | 1.889 |
| SOPS general | Intercept | 8.591 | 0.234 | 36.728 |  |  | Subject | 8.686 | 2.947 |
|  | Baseline tobacco x visit | -0.039 | 0.057 | -0.675 | 0.500 | 1.00 | Residual | 10.025 | 3.166 |
| SAS | Intercept | 44.031 | 0.692 | 63.628 |  |  | Subject | 91.574 | 9.569 |
|  | Baseline tobacco x visit | -0.083 | 0.153 | -0.540 | 0.589 |  | Residual | 65.749 | 8.109 |
| SIAS | Intercept | 30.514 | 0.951 | 32.081 |  |  | Subject | 201.563 | 14.197 |
|  | Baseline tobacco x visit | 0.121 | 0.187 | 0.646 | 0.518 |  | Residual | 95.393 | 9.767 |
| CDSS | Intercept | 5.437 | 0.235 | 23.181 |  |  | Subject | 7.310 | 2.704 |
|  | Baseline tobacco x visit | 0.014 | 0.059 | 0.236 | 0.813 |  | Residual | 10.794 | 3.285 |
| GFR | Intercept | 5.900 | 0.118 | 49.941 |  |  | Subject | 2.658 | 1.630 |
|  | Baseline tobacco x visit | 0.005 | 0.027 | 0.195 | 0.845 |  | Residual | 2.147 | 1.465 |
| GFS | Intercept | 5.879 | 0.084 | 70.291 |  |  | Subject | 1.591 | 1.261 |
|  | Baseline tobacco x visit | -0.033 | 0.017 | -1.969 | **0.049** |  | Residual | 0.822 | 0.907 |
| Global cognition | Intercept | -0.486 | 0.044 | -11.099 |  |  | Subject | 0.502 | 0.708 |
|  | Baseline tobacco x visit | -0.011 | 0.007 | -1.506 | 0.132 |  | Residual | 0.096 | 0.310 |
| Processing speed | Intercept | -0.723 | 0.065 | -11.172 |  |  | Subject | 0.870 | 0.933 |
|  | Baseline tobacco x visit | 0.009 | 0.014 | 0.664 | 0.507 | 1.00 | Residual | 0.382 | 0.618 |
| Working memory | Intercept | -0.531 | 0.065 | -8.188 |  |  | Subject | 0.949 | 0.974 |
|  | Baseline tobacco x visit | -0.033 | 0.013 | -2.620 | 0.009 | 0.061 | Residual | 0.326 | 0.571 |
| Attention | Intercept | -0.624 | 0.064 | -9.703 |  |  | Subject | 0.823 | 0.907 |
|  | Baseline tobacco x visit | -0.025 | 0.014 | -1.851 | 0.064 | 0.385 | Residual | 0.384 | 0.620 |
| Verbal learning | Intercept | -0.584 | 0.073 | -8.002 |  |  | Subject | 0.913 | 0.956 |
|  | Baseline tobacco x visit | 0.002 | 0.017 | 0.110 | 0.913 | 1.00 | Residual | 0.634 | 0.796 |
| Visual learning | Intercept | -0.448 | 0.066 | -6.791 |  |  | Subject | 0.736 | 0.858 |
|  | Baseline tobacco x visit | -0.014 | 0.016 | -0.887 | 0.375 | 1.00 | Residual | 0.523 | 0.723 |
| Problem reasoning | Intercept | -0.115 | 0.062 | -1.855 |  |  | Subject | 0.778 | 0.882 |
|  | Baseline tobacco x visit | 0.013 | 0.013 | 1.007 | 0.314 | 1.00 | Residual | 0.371 | 0.609 |

Cannabis

| Response | Fixed effect |  |  |  |  |  | Random effect |  |  |
| --- | --- | --- | --- | --- | --- | --- | --- | --- | --- |
|  | Term | Estimate | SE | t-value | p-value | FDRp | Term | Variance | SD |
| SOPS total | Intercept | 39.338 | 0.710 | 55.425 |  |  | Subject | 98.759 | 9.938 |
|  | Baseline cannabis x visit | 0.121 | 0.117 | 1.037 | 0.299 |  | Residual | 76.633 | 8.754 |
| SOPS positive | Intercept | 12.217 | 0.231 | 52.895 |  |  | Subject | 10.063 | 3.172 |
|  | Baseline cannabis x visit | -0.023 | 0.039 | -0.592 | 0.554 | 0.554 | Residual | 8.833 | 2.972 |
| SOPS negative | Intercept | 13.146 | 0.321 | 40.908 |  |  | Subject | 21.634 | 4.651 |
|  | Baseline cannabis x visit | 0.159 | 0.051 | 3.145 | **0.0017** | **0.0067** | Residual | 14.303 | 3.782 |
| SOPS disorganization | Intercept | 5.330 | 0.160 | 33.299 |  |  | Subject | 5.360 | 2.315 |
|  | Baseline cannabis x visit | 0.039 | 0.025 | 1.532 | 0.126 | 0.377 | Residual | 3.560 | 1.887 |
| SOPS general | Intercept | 8.673 | 0.232 | 37.363 |  |  | Subject | 8.802 | 2.967 |
|  | Baseline cannabis x visit | -0.048 | 0.042 | -1.142 | 0.254 | 0.507 | Residual | 10.018 | 3.165 |
| SAS | Intercept | 44.437 | 0.689 | 64.520 |  |  | Subject | 92.900 | 9.638 |
|  | Baseline cannabis x visit | -0.027 | 0.110 | -0.241 | 0.891 |  | Residual | 65.784 | 8.111 |
| SIAS | Intercept | 29.972 | 0.943 | 31.789 |  |  | Subject | 201.710 | 14.202 |
|  | Baseline cannabis x visit | -0.063 | 0.134 | -0.468 | 0.639 |  | Residual | 95.421 | 9.768 |
| CDSS | Intercept | 5.392 | 0.232 | 23.205 |  |  | Subject | 7.921 | 2.814 |
|  | Baseline cannabis x visit | -0.066 | 0.043 | -1.515 | 0.130 |  | Residual | 10.721 | 3.274 |
| GFR | Intercept | 5.779 | 0.117 | 49.399 |  |  | Subject | 2.693 | 1.641 |
|  | Baseline cannabis x visit | -0.046 | 0.020 | -2.374 | **0.018** |  | Residual | 2.142 | 1.464 |
| GFS | Intercept | 5.961 | 0.083 | 71.795 |  |  | Subject | 1.613 | 1.270 |
|  | Baseline cannabis x visit | -0.005 | 0.012 | -0.394 | 0.694 |  | Residual | 0.825 | 0.908 |
| Global cognition | Intercept | -0.522 | 0.043 | -12.095 |  |  | Subject | 0.499 | 0.706 |
|  | Baseline cannabis x visit | -0.008 | 0.005 | -1.456 | 0.145 |  | Residual | 0.096 | 0.310 |
| Processing speed | Intercept | -0.751 | 0.064 | -11.753 |  |  | Subject | 0.867 | 0.931 |
|  | Baseline cannabis x visit | 0.006 | 0.010 | 0.609 | 0.423 | 1.000 | Residual | 0.382 | 0.618 |
| Working memory | Intercept | -0.570 | 0.064 | -8.947 |  |  | Subject | 0.926 | 0.962 |
|  | Baseline cannabis x visit | -0.012 | 0.009 | -1.299 | 0.339 | 0.873 | Residual | 0.329 | 0.574 |
| Attention | Intercept | -0.658 | 0.063 | -10.385 |  |  | Subject | 0.818 | 0.904 |
|  | Baseline cannabis x visit | -0.015 | 0.010 | -1.435 | 0.226 | 0.873 | Residual | 0.386 | 0.621 |
| Verbal learning | Intercept | -0.607 | 0.072 | -8.417 |  |  | Subject | 0.914 | 0.956 |
|  | Baseline cannabis x visit | 0.005 | 0.013 | 0.414 | 0.729 | 1.000 | Residual | 0.635 | 0.797 |
| Visual learning | Intercept | -0.520 | 0.065 | -7.987 |  |  | Subject | 0.739 | 0.859 |
|  | Baseline cannabis x visit | -0.031 | 0.011 | -2.720 | **0.00657** | **0.046** | Residual | 0.518 | 0.720 |
| Problem reasoning | Intercept | -0.149 | 0.061 | -2.426 |  |  | Subject | 0.778 | 0.882 |
|  | Baseline cannabis x visit | 0.011 | 0.010 | 1.101 | 0.206 | 0.873 | Residual | 0.371 | 0.609 |
